# Validation of 13,102 ICD-10-CM Codes Using a Large Language Model-Based System

**DOI:** 10.64898/2025.12.30.25343244

**Authors:** Yichen Wang, Yilin Song, Rex Siu, Induja R. Nimma, Yan Yan, Thomas R. Savage, Yiming Wang, Zhichen Li, Daryl Ramai, Jiale Wang, Dilhana Badurdeen, Cui Tao, Vivek Kumbhari, Yuting Huang

## Abstract

**Objective:** To comprehensively evaluate the validity of ICD-10-CM codes for both prevalent diagnoses and less common diseases, and to assess the performance of a large language model (LLM)-based system in validating these codes.

**Materials and Methods:** This retrospective study analyzed hospital admissions from the Medical Information Mart for Intensive Care (MIMIC-IV) database. We developed a validated LLM-based system using GPT-4o, refined through iterative prompt engineering, to assess ICD-10-CM code validity. We measured the PPV of ICD-10-CM codes, PPV of principal and secondary diagnoses, and the performance of an LLM-based system in code validation.

**Results:** Among 865,079 assigned codes, the PPV was 84.6% (95% CI, 84.5%-84.6%). Principal diagnoses had a PPV of 93.9% (95% CI, 93.7%-94.1%), while secondary diagnoses had a PPV of 83.8% (95% CI, 83.7%-83.9%). The LLM system demonstrated high performance in validating ICD codes, achieving 93.6% accuracy, 95.4% sensitivity and 85.2% specificity. Among correctly assigned secondary diagnoses, the majority (67.9%) represented historical or baseline conditions, while 32.1% reflected active conditions that deviated from baseline status; 22.3% of these emerged after hospital admission. PPV decreases with later diagnosis positions, with the largest decline occurring between principal and secondary diagnoses.

**Discussion and Conclusion:** In this large-scale evaluation, ICD-10-CM codes exhibited generally high accuracy, though variability existed by position and condition type. A validated LLM system performed comparably to physician review and offers a scalable means to improve coding accuracy. These findings support the potential for integrating LLM-based auditing into routine workflows to strengthen the quality of administrative and research data.

## INTRODUCTION

The International Classification of Diseases, Tenth Revision (ICD-10) is extensively used worldwide to classify diagnoses for billing, outcomes research, epidemiology, and policy decisions, with the World Health Organization estimating that over 100 countries rely on ICD-10 for healthcare claims and mortality statistics.^1,2^ In the U.S. alone, nearly all of the nation’s healthcare expenditures (exceeding $4.5 trillion in 2022) flow through reimbursement processes requiring ICD-10-CM codes, which drives an annual volume of more than three billion claims.^3,4^ Moreover, tens of thousands of publications reference ICD-10, reflecting its indispensable role in large-scale population health studies, disease surveillance, and outcome research.^5^ Health insurers increasingly incorporate ICD-10 codes into risk-adjustment models, while policymakers use coded data to guide public health interventions and resource allocation.^1^

Although some studies have evaluated the validity of specific codes or small subsets of codes,^6–8^ few have investigated a broad spectrum at scale. Most prior efforts relied on labor-intensive manual chart review, which limits reproducibility and scalability. These studies typically focused on a narrow set of conditions with limited generalizability to the broader ICD-10 classification system. Moreover, the proportion of codes that accurately reflect active clinical problems remains unclear.^9^ Recent advancements in large language models (LLMs) that can handle large volumes of data quickly and consistently offer an exciting way to address these challenges.^10,11^

Manual coding workflows remain error-prone, with published validity estimates for ICD-10 codes ranging from 53% to 98% across different conditions and healthcare settings.^12,13^ Systematic reviews have reported discharge coding inaccuracies in 10%–20% of cases.^13^ Against this backdrop, scalable LLM-based auditing tools have the potential to substantially improve coding validity and reduce administrative burden.

In this study, we take a comprehensive look at real-world ICD-10-CM code accuracy. We apply a validated GPT-4o-based system to examine 13,102 codes documented across 64,764 hospital admissions. Our aim is to highlight the current state of ICD-10-CM coding accuracy and show how an LLM can support large-scale code validation.

We define real-world ICD-10-CM code “accuracy” as positive predictive value (PPV) – the probability that a patient truly has a condition when the corresponding ICD-10-CM code is present. This measure is more appropriate than traditional statistical “accuracy,” which includes true negatives (patients without the code and without the condition). In real-world datasets, true negatives constitute the vast majority, often inflating overall accuracy even when code performance is poor. However, while PPV reflects the likelihood that coded cases are true cases, it does not capture sensitivity – that is, the proportion of all true cases that are coded – thus providing no insight into undercoding.

To evaluate the performance of the LLM in determining the correctness of ICD-10-CM code assignments, we use standard classification metrics: accuracy (in its traditional statistical sense), sensitivity, specificity, PPV, and negative predictive value (NPV).

## METHODS

### Data Collection and Variables

We used the Medical Information Mart for Intensive Care (MIMIC-IV) database, a publicly available, de-identified electronic health record dataset.^14^ MIMIC-IV includes comprehensive clinical data from patients admitted to a tertiary teaching hospital in Boston, Massachusetts, between 2008 and 2019. We randomly selected 70,000 hospital admissions from 2016 to 2019. 5,236 records could not be successfully processed due to exceeding the model’s maximum context window or failing to produce the required structured output format, despite automated retry mechanisms. These records were excluded from analysis, resulting in a final analytic cohort of 64,764 admissions.

Full-length discharge summaries, ICD-10-CM codes, and corresponding standard code descriptions were provided to the LLM. Further information provided included demographic variables such as age, sex, and race/ethnicity, body mass index, laboratory test results, and vital signs. The dataset encompassed the complete range of recorded minimum and maximum values for laboratory tests and vital signs during hospitalization. Microbiology culture results, including antibiotic susceptibility interpretations, were also included. Furthermore, additional clinical parameters, such as fluid balance calculations, urine output, and device-related metrics (e.g., ventilator settings, central venous pressure), were provided if available.

Additional variables provided contextual insights and served as potential predictors of ICD code misclassification, including insurance type, primary language, admission type (emergency, observation, direct admission, or surgical same-day), admission source (physician referral, emergency room, inter-hospital transfer), length of hospital stays, need for intensive car unit (ICU) admission, discharge disposition, and in-hospital mortality. This study was deemed exempt by the Mayo Clinic Florida Institutional Review Board.

### Outcomes

We defined three outcome measures to evaluate ICD-10-CM code quality. First, we assessed the overall PPV of ICD code assignments, defined as the proportion of assigned codes that accurately reflected conditions documented in the medical record. Second, among correctly assigned secondary diagnoses, we evaluated clinical relevance by determining the PPV for active conditions, defined as diseases that reflect deviation from patients’ baseline health status (e.g., new onset or acute exacerbation of chronic disease). Active conditions were defined as medical issues that represented a clinically meaningful change from baseline, such as acute illnesses or subacute worsening of chronic conditions, and were determined based on LLM review of the discharge summary in the context of the patient’s hospital course. Stable or historical conditions, in contrast, were those present at baseline or unrelated to the index admission. This distinction was made by the LLM based on a structured prompt, which guided classification independent of code terminology. Third, for active conditions, we calculated the PPV for post-admission onset, defined as conditions that developed after hospital admission rather than being present on arrival. Classification of activeness and post-admission onset was performed via a separate LLM query and constituted a secondary outcome. The prompts used for all tasks are provided in the **Supplementary Materials**.

### LLM System Configuration

We utilized GPT-4o through Mayo Clinic Cloud, an institutional access, to develop our LLM system. The system was built using a systematic prompt engineering approach that underwent multiple iterations of refinement. The prompt template was modified based on error analysis of the LLM’s outputs until satisfactory alignment with a physician developer was achieved. Approximately 50 discharge summaries were used during this development phase. A physician prompt developer manually reviewed model responses and iteratively refined the prompt to clarify ambiguous instructions, address known sources of misunderstanding, and incorporate reasoning steps to minimize common diagnostic errors. Chain-of-thought prompting was employed to encourage structured clinical reasoning. The final prompt was considered validated after achieving consistent agreement with physician review across 10 consecutive discharge summaries.

The LLM system was designed to assess the validity of each code based on supporting documentation, adherence to diagnostic criteria, and reflect on common diagnostic errors among trainees. To enhance reliability and minimize hallucinations, each chart was independently reviewed by the LLM three times. Code validity was determined by majority vote – considered correct if at least two of three evaluations concurred. Detailed system parameters and an example LLM response are provided in the supplementary materials.

### Evaluation of the LLM System’s Performance

We conducted a structured pre-deployment evaluation to assess the LLM system’s performance. In this phase, two physicians independently reviewed a sample set of codes. Any discrepancies were resolved through arbitration by a third physician, thereby establishing the physician panel’s consensus. In cases of disagreement between the physician panel and the LLM, the underlying medical record was re-reviewed by physicians. If the LLM highlighted supporting clinical evidence that had been initially overlooked, and this evidence was confirmed on physician re-review, the reconciled physician-LLM judgment was incorporated into the reference standard. Importantly, final adjudication was determined by physician confirmation rather than by the LLM alone. This allowed us to leverage the LLM’s consistency and attention to detail while maintaining physician oversight for contextual interpretation and final adjudication. However, we recognize that involving the LLM in constructing the reference standard may introduce potential bias. To address potential bias introduced by this adjudication approach, we conducted a sensitivity analysis using a strictly physician-only reference standard, in which LLM judgments were not used in determining code correctness. In this secondary evaluation, two physicians independently reviewed 10 randomly selected discharge summaries (175 ICD-10-CM codes), and a third adjudicated disagreements. The LLM’s performance was then compared against this entirely physician-derived standard to assess potential bias in our original evaluation process.

After completion of the pre-deployment analysis, the LLM system was deployed across a cohort of 70,000 hospital admissions. A post-deployment evaluation was subsequently performed by randomly sampling the LLM-generated responses for secondary review. To optimize the number of cases reviewed, each response was evaluated by a single physician; if disagreement with the LLM’s judgment arose, a second physician conducted a follow-up review. **Figure 1** summarizes the study design.

**Figure 1.**
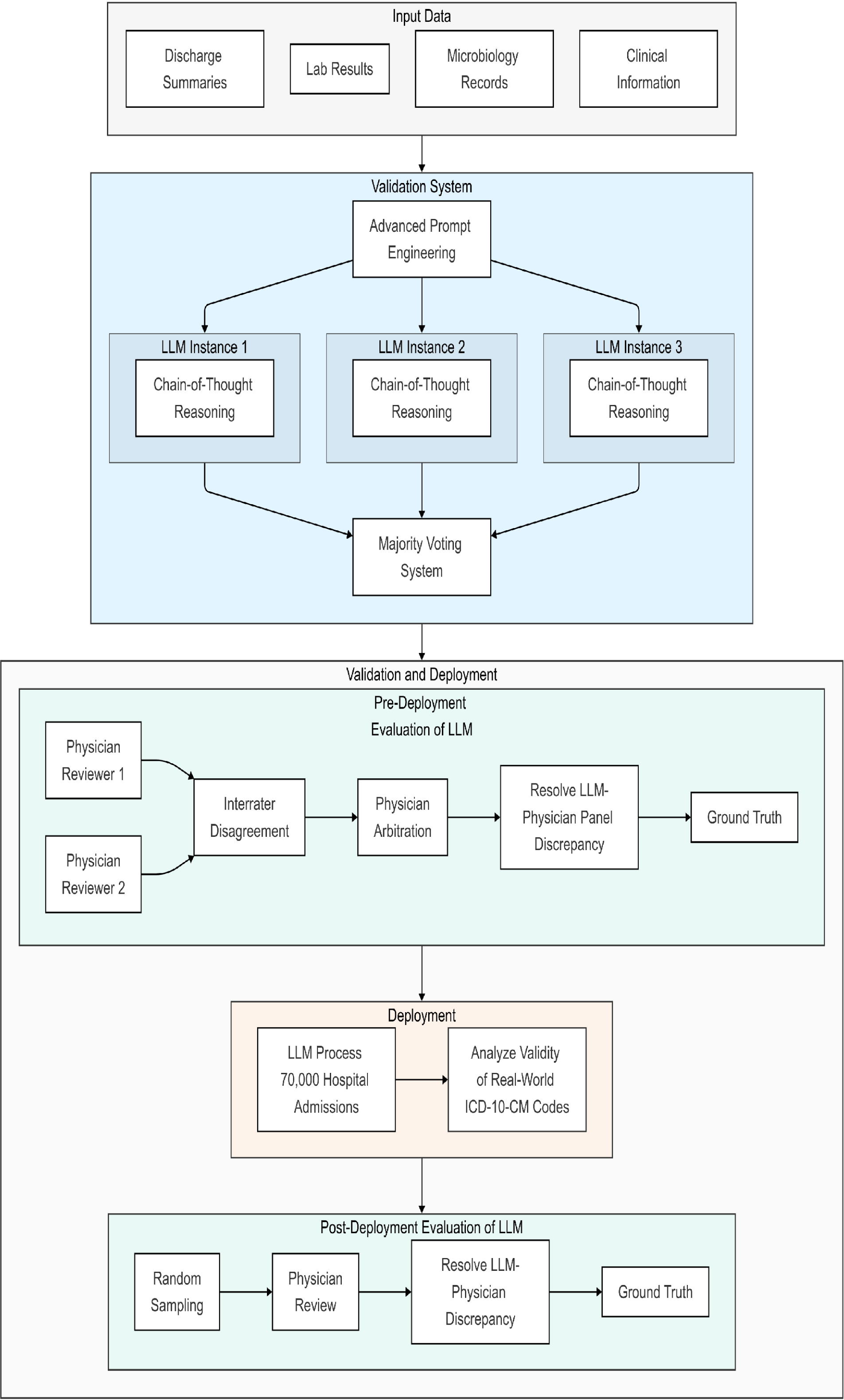
System Architecture and Two-Phase Evaluation Process for ICD-10-CM Code Validation Using Large Language Models. This figure illustrates the systematic approach for validating ICD-10-CM codes using a large language model (LLM)-based system. The validation system incorporates multiple data sources and employs three parallel LLM instances with chain-of-thought reasoning and majority voting. The evaluation process consists of three sequential phases: (1) pre-deployment evaluation of LLM system performance involving dual physician review with interrater assessment; (2) large-scale processing of 70,000 hospital admissions and subsequent analysis of real-world ICD-10-CM’s validity; and (3) post-deployment evaluation through random sampling and independent physician review to validate LLM system performance. The system was deployed across 70,000 hospital admissions, of which 64,764 were successfully processed and included in the final analysis.

### Statistical Analysis

All analyses were conducted using Stata 17.0 (StataCorp LLC, College Station, TX) and Python 3.11.10 (Python Software Foundation, Wilmington, DE). Continuous variables were reported as medians with interquartile ranges (IQR) due to their non-normal distributions, while categorical variables (e.g., sex, race, insurance type) were reported as counts and percentages.

To assess the performance of the LLM system in validating ICD-10-CM codes, we computed sensitivity, specificity, PPV, and NPV. These metrics were further stratified based on ICD-10-CM code frequency to evaluate performance consistency across common and rare codes. To account for potential bias due to insufficient documentation, we performed a sensitivity analysis excluding codes in the lowest quartile of PPV.

Logistic regression models were employed to identify predictors of incorrect ICD-10-CM assignments and LLM judgments. To assess the impact of diagnosis position on coding accuracy, we conducted a linear regression analysis, modeling PPV as a function of diagnosis position. Statistical significance was set at a two-sided p-value <0.05.

## RESULTS

### Patient and Hospitalization Characteristics

There were 64,764 admissions included in this study. The median age was 61 years and 50.8% were female. The study population was predominantly White (67.3%), followed by Black (15.0%), Hispanic (5.0%), and Asian (3.8%) patients. Medicare was the most common insurance type (39.0%), while 5.8% had Medicaid coverage. English was the primary language for 89.4% of patients. Nearly half of admissions were for observation status (49.6%). During hospitalization, 19.6% required ICU-level care, and the median length of stay was 4 days (IQR: 2-6). Most patients were discharged home (74.6%). The in-hospital mortality rate was 2.3%. Detailed patient and hospitalization characteristics can be found in **Table 1**.

**Table 1.**
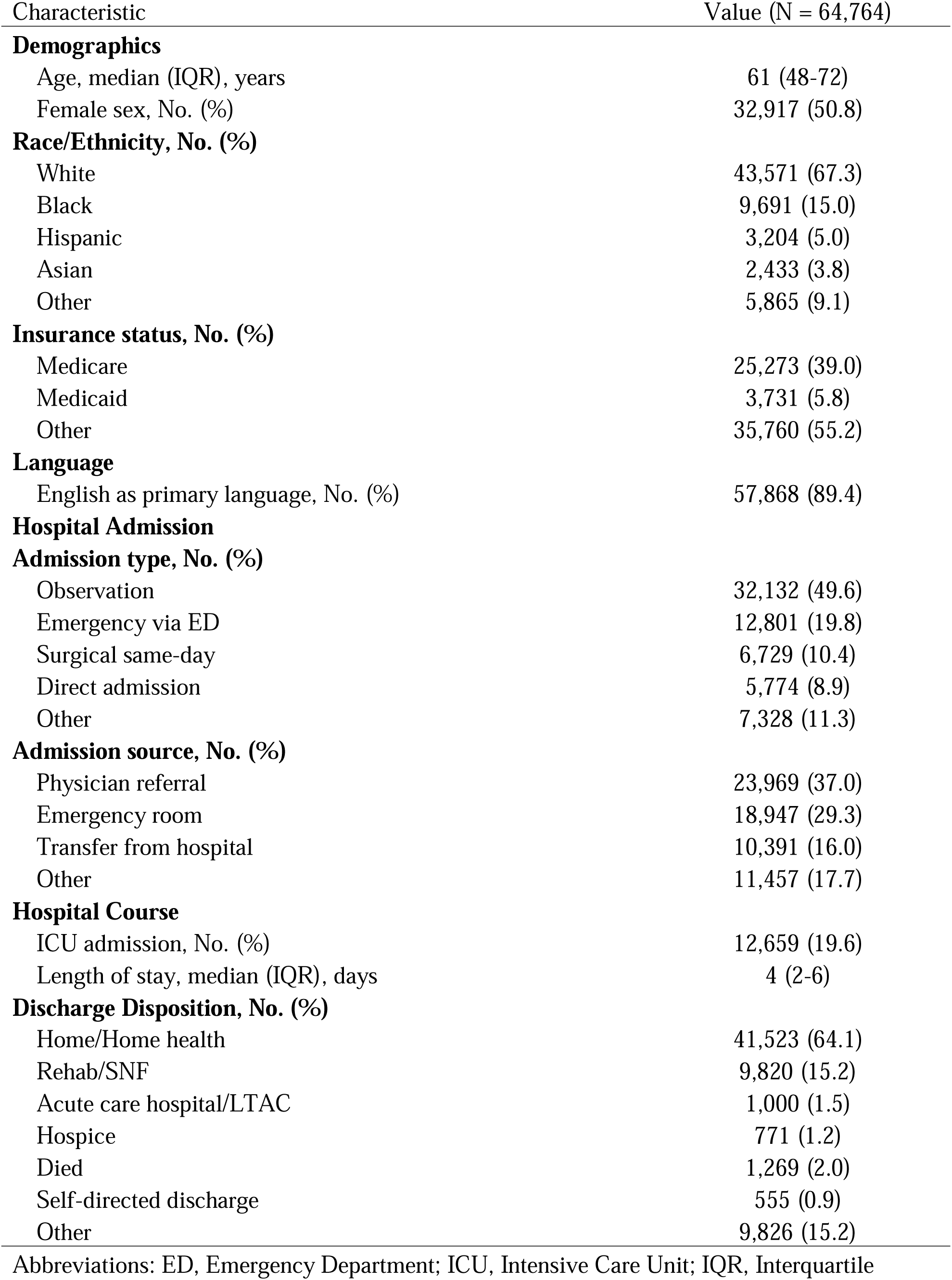
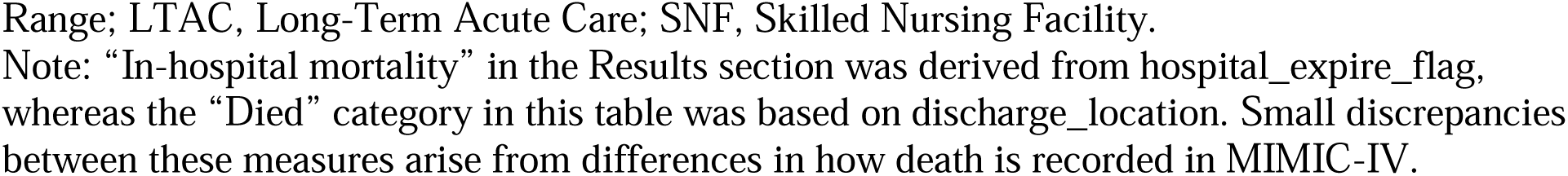
Baseline Characteristics of Study Population.

| Characteristic | Value (N = 64,764) |
| --- | --- |
| **Demographics** |  |
| Age, median (IQR), years | 61 (48-72) |
| Female sex, No. (%) | 32,917 (50.8) |
| **Race/Ethnicity, No. (%)** |  |
| White | 43,571 (67.3) |
| Black | 9,691 (15.0) |
| Hispanic | 3,204 (5.0) |
| Asian | 2,433 (3.8) |
| Other | 5,865 (9.1) |
| **Insurance status, No. (%)** |  |
| Medicare | 25,273 (39.0) |
| Medicaid | 3,731 (5.8) |
| Other | 35,760 (55.2) |
| **Language** |  |
| English as primary language, No. (%) | 57,868 (89.4) |
| **Hospital Admission** |  |
| **Admission type, No. (%)** |  |
| Observation | 32,132 (49.6) |
| Emergency via ED | 12,801 (19.8) |
| Surgical same-day | 6,729 (10.4) |
| Direct admission | 5,774 (8.9) |
| Other | 7,328 (11.3) |
| **Admission source, No. (%)** |  |
| Physician referral | 23,969 (37.0) |
| Emergency room | 18,947 (29.3) |
| Transfer from hospital | 10,391 (16.0) |
| Other | 11,457 (17.7) |
| **Hospital Course** |  |
| ICU admission, No. (%) | 12,659 (19.6) |
| Length of stay, median (IQR), days | 4 (2-6) |
| **Discharge Disposition, No. (%)** |  |
| Home/Home health | 41,523 (64.1) |
| Rehab/SNF | 9,820 (15.2) |
| Acute care hospital/LTAC | 1,000 (1.5) |
| Hospice | 771 (1.2) |
| Died | 1,269 (2.0) |
| Self-directed discharge | 555 (0.9) |
| Other | 9,826 (15.2) |

Abbreviations: ED, Emergency Department; ICU, Intensive Care Unit; IQR, Interquartile
Range; LTAC, Long-Term Acute Care; SNF, Skilled Nursing Facility.
Note: “In-hospital mortality” in the Results section was derived from hospital\_expire\_flag, whereas the “Died” category in this table was based on discharge\_location. Small discrepancies between these measures arise from differences in how death is recorded in MIMIC-IV.

### Validation of the LLM System

In pre-deployment analysis (N=295), two physician reviewers achieved correct code assessments in 90.5% and 87.5% of cases, compared with 92.2% for the physician panel and 93.6% for the LLM system. The accuracy difference between the panel and the LLM was not statistically significant (P=0.522). Cohen κ was 0.76 between the two individual physicians and 0.51 between the panel and the LLM, indicating substantial and moderate agreement, respectively. Against the ground truth, the LLM showed the highest sensitivity (95.4%), specificity (85.2%), PPV (96.6%), and NPV (80.7%) (**Figure 2** and **Figure 3**). Among all ICD-10 code assignment, 80.7% were truly correct per ground truth; the LLM predicted 81.7% correct and the physician panel 83.1% correct.

**Figure 2.**
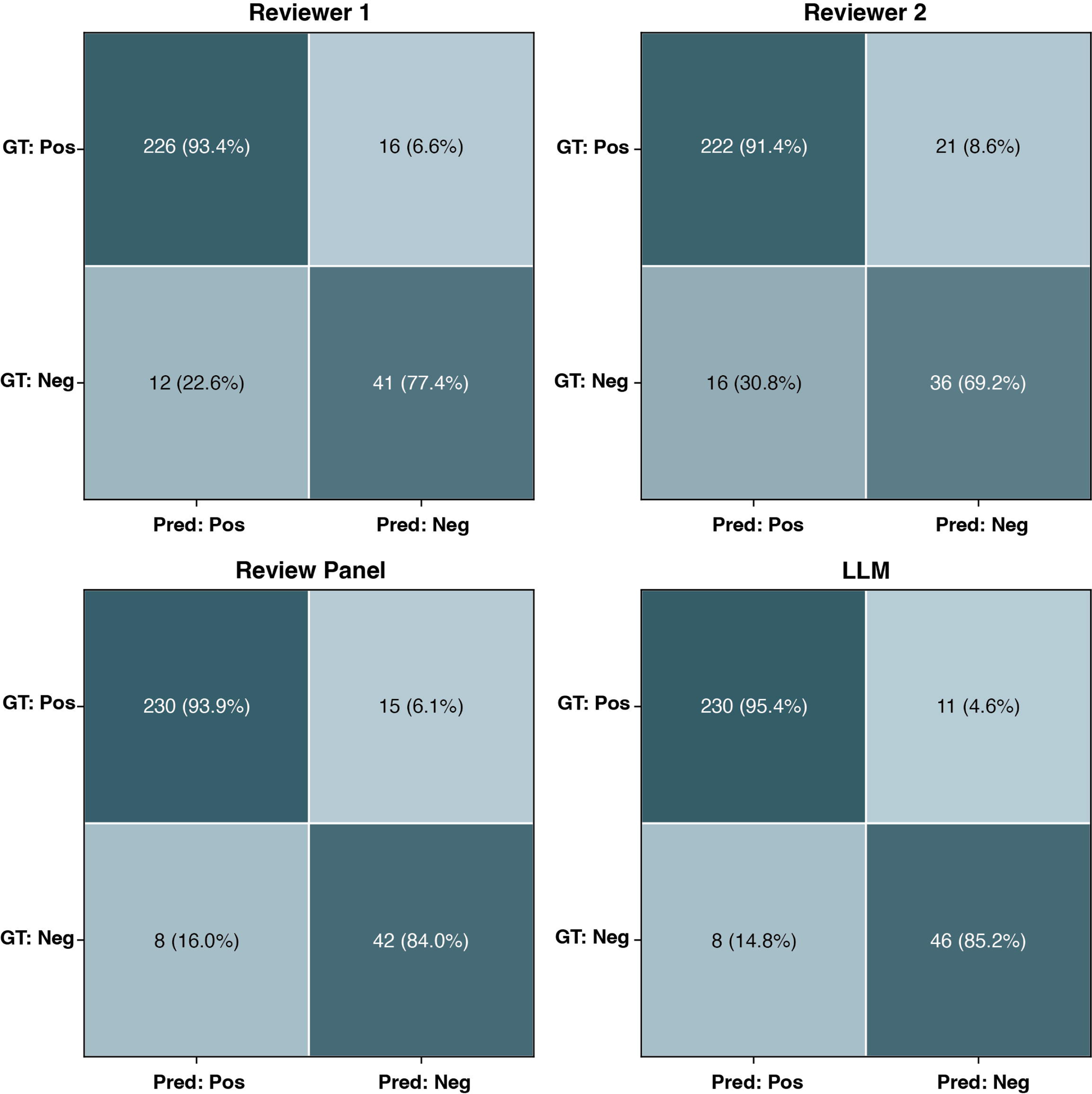
Comparison LLM System and Human on Performance on ICD-10 Validation Tasks. This bar chart compares five key diagnostic performance metrics – accuracy, sensitivity, specificity, positive predictive value (PPV), and negative predictive value (NPV) – for LLM system and human evaluations.

**Figure 3.**
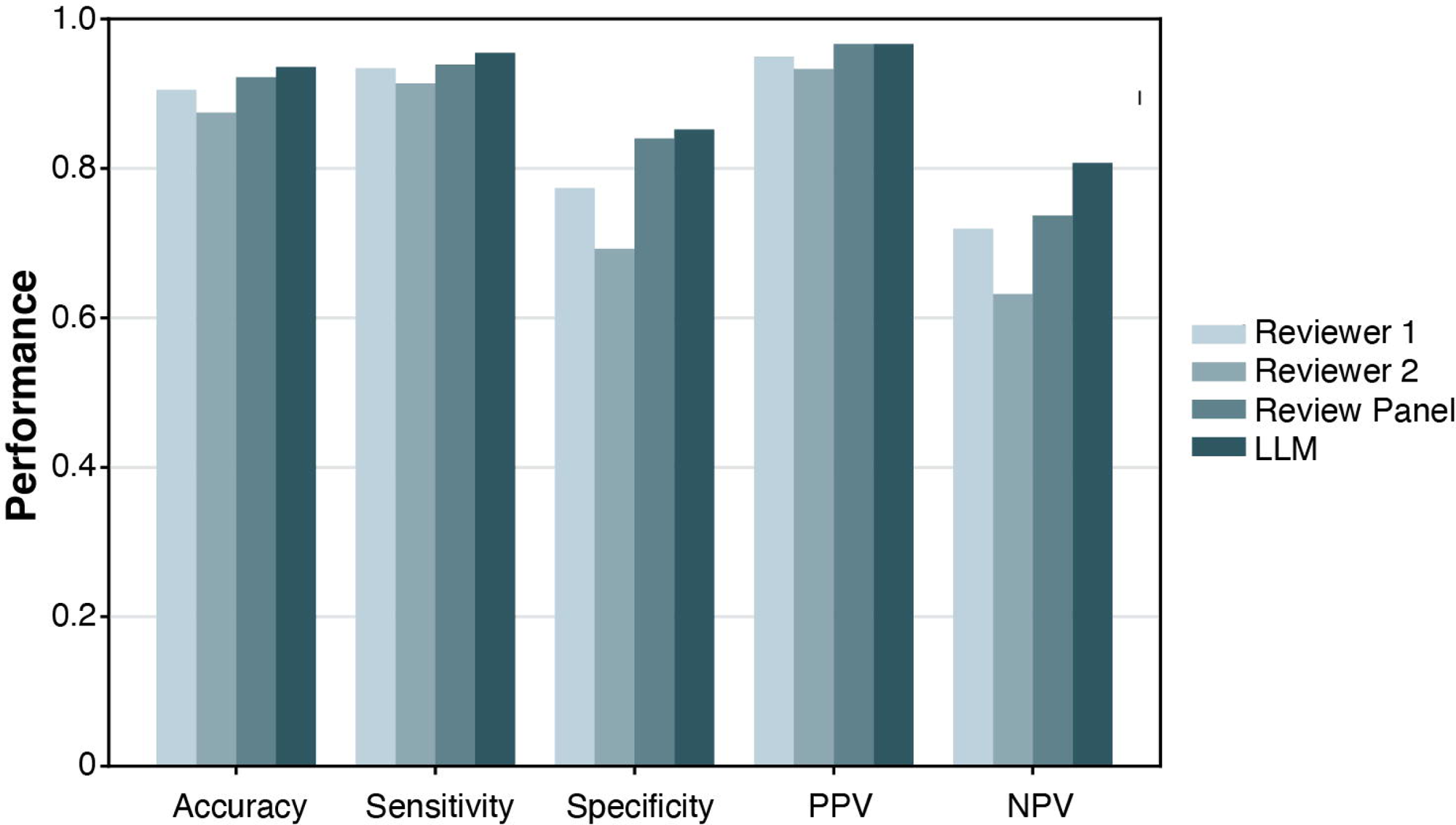
Confusion Matrices for LLM System and Human Predictions. These heatmaps present the confusion matrices for LLM system and human reviewer predictions. For each matrix, rows represent the ground truth (positive for correct ICD-10 assignment, negative for incorrect ICD-10 assignment) and columns represent the LLM system’s or human’s judgment on the correctness of ICD-10 assignments. Each cell displays both the raw frequency (number of cases) and the row-normalized rate (percentage). The cell colors are based on the rate (ranging from 0% to 100%), using a light-to-dark color gradient to facilitate visual comparison.

In the independent sensitivity analysis using a physician-only reference standard (10 discharge summaries, 175 codes), the LLM demonstrated a sensitivity of 94.2%, specificity of 80.0%, PPV of 97.3%, and NPV of 64.0%.

In post-deployment analysis (N=971), the LLM system achieved accuracy (97.8%), sensitivity (98.5%), and specificity (94.5%), with PPV and NPV of 98.9% and 92.8%, respectively. Performance remained consistent across code frequencies (**Supplementary Table S1**). For the top 10% most frequent codes (N=429; 3–33 occurrences), sensitivity and specificity were 97.7% and 95.2%. For the bottom 10% least frequent codes (N=386; single occurrences), sensitivity was 99.1%, specificity 93.4%. The area under the receiver operating characteristic curve remained stable at 0.96 across all subgroups, indicating consistent discriminative ability.

Logistic regression (**Supplementary Table S2**) found no significant predictors of LLM errors across demographics, clinical features, or admission variables. Patient factors (e.g., age: OR 1.00, 95% CI 0.97–1.03; female sex: OR 0.43, 95% CI 0.14–1.29), clinical complexity (e.g., number of diagnoses: OR 1.06, 95% CI 0.99–1.13), and admission type (e.g., emergency vs. observation: OR 1.87, 95% CI 0.67–5.21) showed no association, suggesting stable system performance across patient and administrative contexts.

### Validation of ICD-10-CM Code

Using our validated LLM system, we analyzed 865,079 ICD-10-CM code assignments (13,102 distinct codes) across 64,764 hospital admissions. The overall PPV of ICD-10-CM codes was 84.6% (95% CI 84.5% to 84.6%). Principal diagnoses demonstrated the highest PPV at 93.9% (95% CI 93.7% to 94.1%), while secondary diagnoses showed a lower PPV of 83.8% (95% CI 83.7% to 83.9%). Across individual ICD-10-CM codes, the median PPV was 92.3% (IQR: 66.7%-100%). The distribution of PPV was left-skewed, indicating the presence of outliers with particularly low PPVs (**Supplementary Figure S1**). A comprehensive breakdown of PPV by ICD code frequency is provided in **Table 2** and available at the online repository.

**Table 2.**
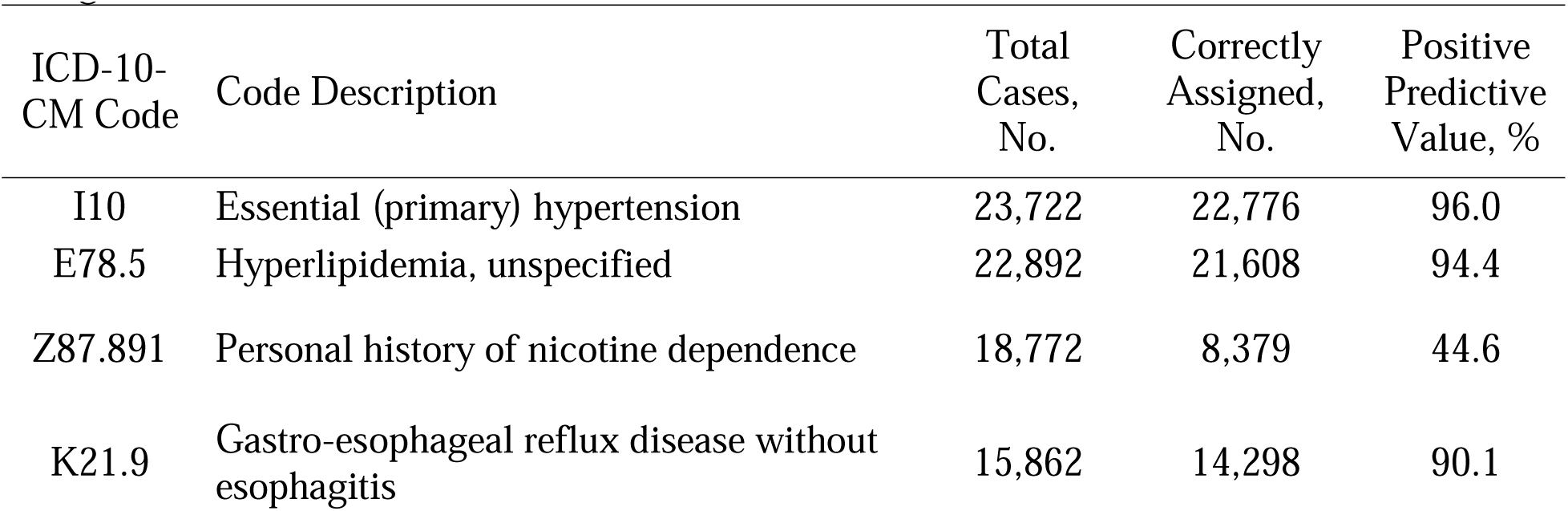
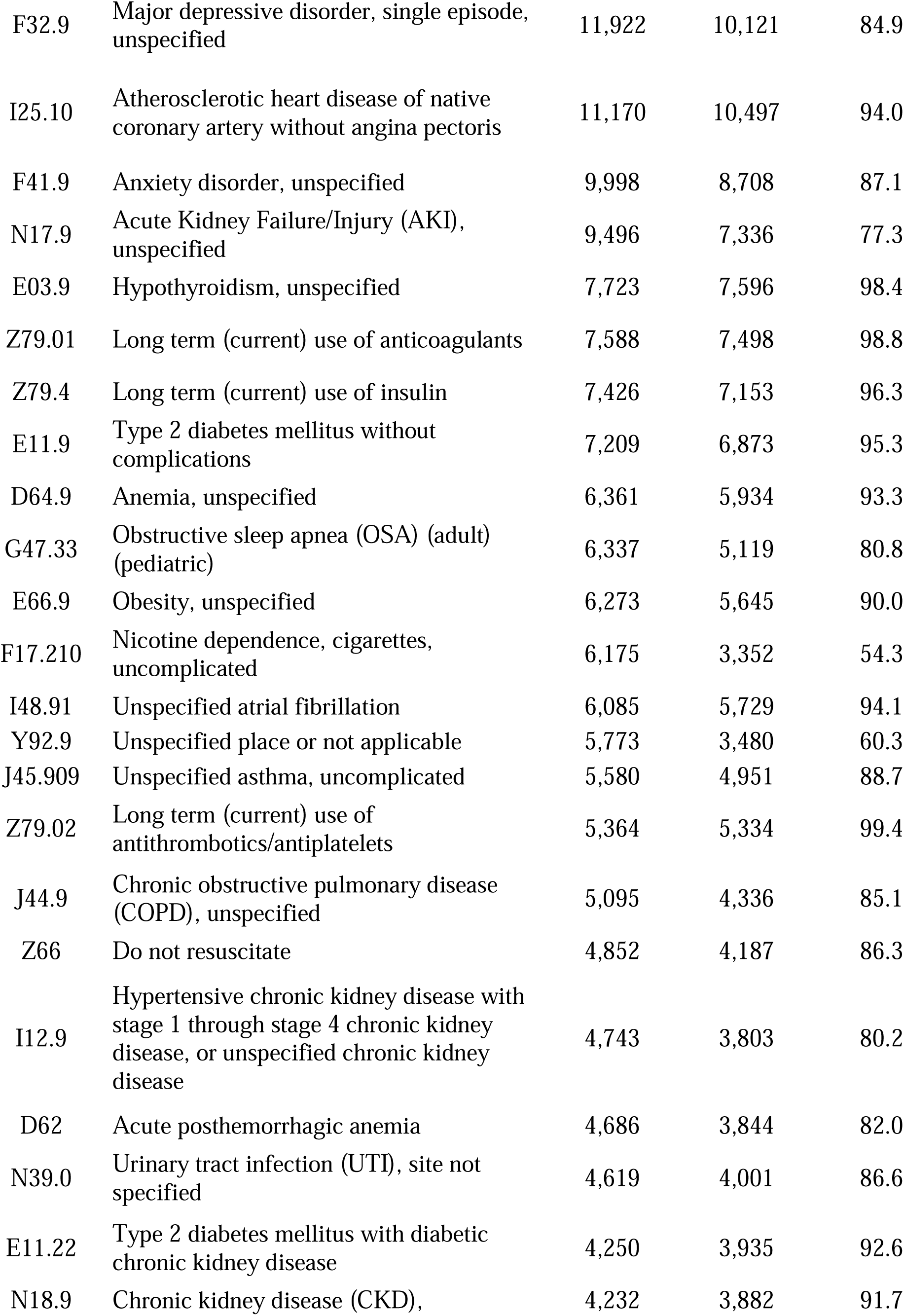
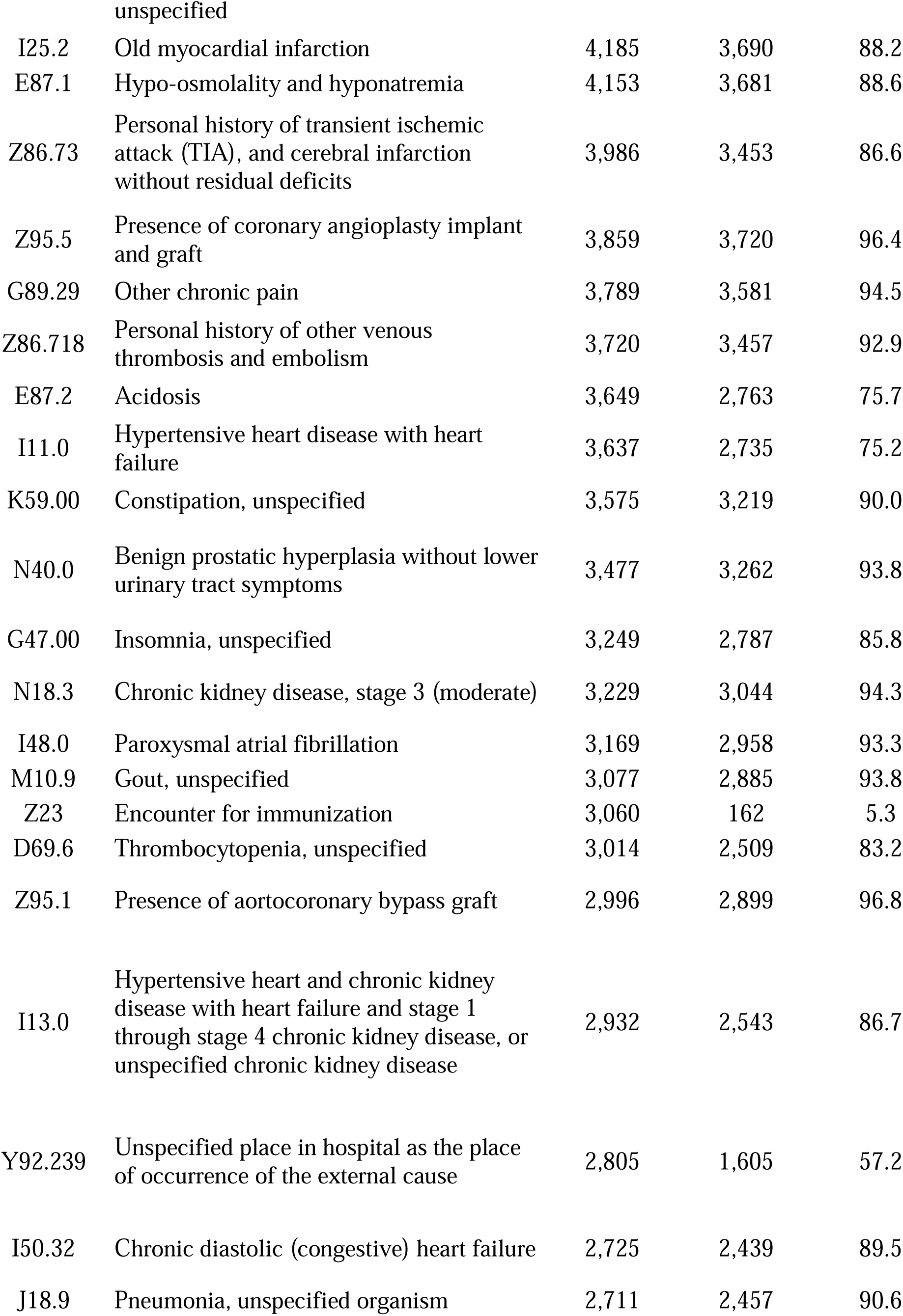
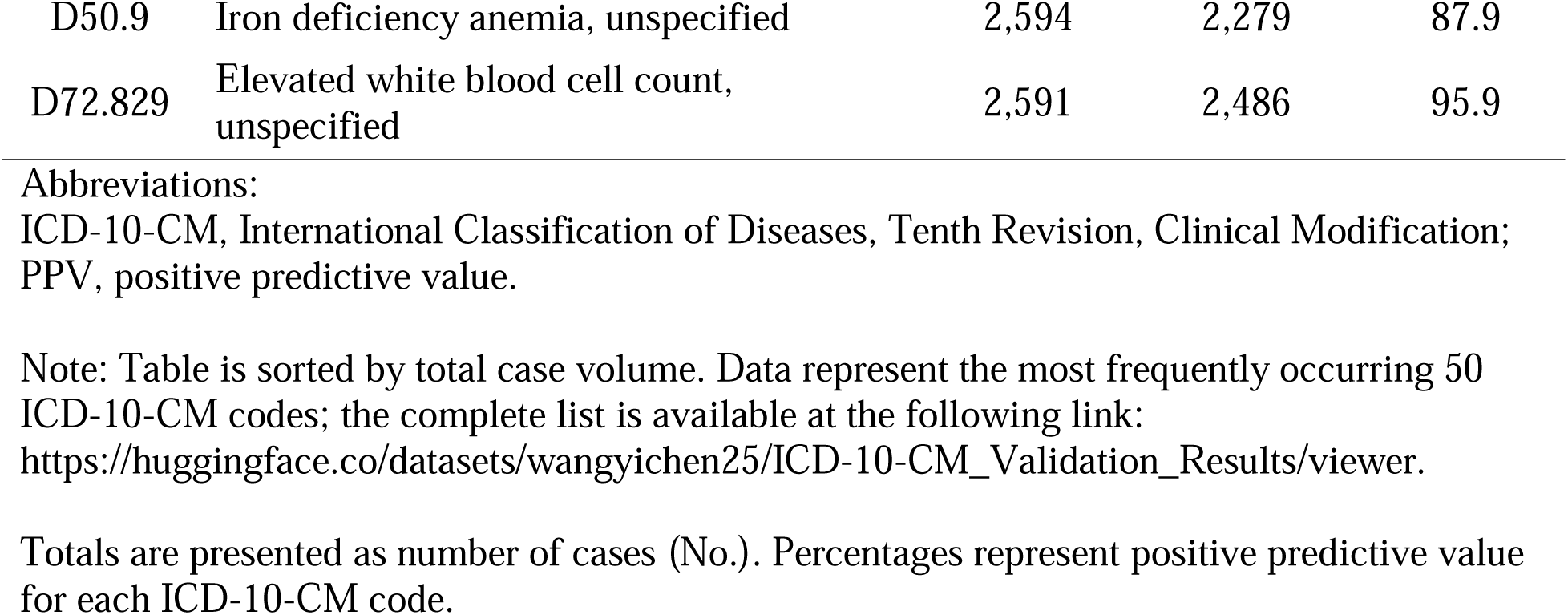
Positive Predictive Value of the 50 Most Frequently Occurring ICD-10-CM Diagnoses.

| ICD-10-CM Code | Code Description | Total Cases, No. | Correctly Assigned, No. | Positive Predictive Value, % |
| --- | --- | --- | --- | --- |
| I10 | Essential (primary) hypertension | 23,722 | 22,776 | 96.0 |
| E78.5 | Hyperlipidemia, unspecified | 22,892 | 21,608 | 94.4 |
| Z87.891 | Personal history of nicotine dependence | 18,772 | 8,379 | 44.6 |
| K21.9 | Gastro-esophageal reflux disease without esophagitis | 15,862 | 14,298 | 90.1 |
| F32.9 | Major depressive disorder, single episode, unspecified | 11,922 | 10,121 | 84.9 |
| I25.10 | Atherosclerotic heart disease of native coronary artery without angina pectoris | 11,170 | 10,497 | 94.0 |
| F41.9 | Anxiety disorder, unspecified | 9,998 | 8,708 | 87.1 |
| N17.9 | Acute Kidney Failure/Injury (AKI), unspecified | 9,496 | 7,336 | 77.3 |
| E03.9 | Hypothyroidism, unspecified | 7,723 | 7,596 | 98.4 |
| Z79.01 | Long term (current) use of anticoagulants | 7,588 | 7,498 | 98.8 |
| Z79.4 | Long term (current) use of insulin | 7,426 | 7,153 | 96.3 |
| E11.9 | Type 2 diabetes mellitus without complications | 7,209 | 6,873 | 95.3 |
| D64.9 | Anemia, unspecified | 6,361 | 5,934 | 93.3 |
| G47.33 | Obstructive sleep apnea (OSA) (adult) (pediatric) | 6,337 | 5,119 | 80.8 |
| E66.9 | Obesity, unspecified | 6,273 | 5,645 | 90.0 |
| F17.210 | Nicotine dependence, cigarettes, uncomplicated | 6,175 | 3,352 | 54.3 |
| I48.91 | Unspecified atrial fibrillation | 6,085 | 5,729 | 94.1 |
| Y92.9 | Unspecified place or not applicable | 5,773 | 3,480 | 60.3 |
| J45.909 | Unspecified asthma, uncomplicated | 5,580 | 4,951 | 88.7 |
| Z79.02 | Long term (current) use of antithrombotics/antiplatelets | 5,364 | 5,334 | 99.4 |
| J44.9 | Chronic obstructive pulmonary disease (COPD), unspecified | 5,095 | 4,336 | 85.1 |
| Z66 | Do not resuscitate | 4,852 | 4,187 | 86.3 |
| I12.9 | Hypertensive chronic kidney disease with stage 1 through stage 4 chronic kidney disease, or unspecified chronic kidney disease | 4,743 | 3,803 | 80.2 |
| D62 | Acute posthemorrhagic anemia | 4,686 | 3,844 | 82.0 |
| N39.0 | Urinary tract infection (UTI), site not specified | 4,619 | 4,001 | 86.6 |
| E11.22 | Type 2 diabetes mellitus with diabetic chronic kidney disease | 4,250 | 3,935 | 92.6 |
| N18.9 | Chronic kidney disease (CKD), unspecified | 4,232 | 3,882 | 91.7 |
| I25.2 | Old myocardial infarction | 4,185 | 3,690 | 88.2 |
| E87.1 | Hypo-osmolality and hyponatremia | 4,153 | 3,681 | 88.6 |
| Z86.73 | Personal history of transient ischemic attack (TIA), and cerebral infarction without residual deficits | 3,986 | 3,453 | 86.6 |
| Z95.5 | Presence of coronary angioplasty implant and graft | 3,859 | 3,720 | 96.4 |
| G89.29 | Other chronic pain | 3,789 | 3,581 | 94.5 |
| Z86.718 | Personal history of other venous thrombosis and embolism | 3,720 | 3,457 | 92.9 |
| E87.2 | Acidosis | 3,649 | 2,763 | 75.7 |
| I11.0 | Hypertensive heart disease with heart failure | 3,637 | 2,735 | 75.2 |
| K59.00 | Constipation, unspecified | 3,575 | 3,219 | 90.0 |
| N40.0 | Benign prostatic hyperplasia without lower urinary tract symptoms | 3,477 | 3,262 | 93.8 |
| G47.00 | Insomnia, unspecified | 3,249 | 2,787 | 85.8 |
| N18.3 | Chronic kidney disease, stage 3 (moderate) | 3,229 | 3,044 | 94.3 |
| I48.0 | Paroxysmal atrial fibrillation | 3,169 | 2,958 | 93.3 |
| M10.9 | Gout, unspecified | 3,077 | 2,885 | 93.8 |
| Z23 | Encounter for immunization | 3,060 | 162 | 5.3 |
| D69.6 | Thrombocytopenia, unspecified | 3,014 | 2,509 | 83.2 |
| Z95.1 | Presence of aortocoronary bypass graft | 2,996 | 2,899 | 96.8 |
| I13.0 | Hypertensive heart and chronic kidney disease with heart failure and stage 1 through stage 4 chronic kidney disease, or unspecified chronic kidney disease | 2,932 | 2,543 | 86.7 |
| Y92.239 | Unspecified place in hospital as the place of occurrence of the external cause | 2,805 | 1,605 | 57.2 |
| I50.32 | Chronic diastolic (congestive) heart failure | 2,725 | 2,439 | 89.5 |
| J18.9 | Pneumonia, unspecified organism | 2,711 | 2,457 | 90.6 |
| D50.9 | Iron deficiency anemia, unspecified | 2,594 | 2,279 | 87.9 |
| D72.829 | Elevated white blood cell count, unspecified | 2,591 | 2,486 | 95.9 |

Abbreviations:
ICD-10-CM, International Classification of Diseases, Tenth Revision, Clinical Modification; PPV, positive predictive value.
Note: Table is sorted by total case volume. Data represent the most frequently occurring 50 ICD-10-CM codes; the complete list is available at the following link:
https://huggingface.co/datasets/wangyichen25/ICD-10-CM_Validation_Results/viewer.
Totals are presented as number of cases (No.). Percentages represent positive predictive value for each ICD-10-CM code.

A sensitivity analysis excluding codes in the lowest PPV quartile (0–66.7%; e.g., “encounter for immunization”) yielded an increased overall PPV of 91.5% (95% CI, 91.4%–91.5%), with principal and secondary diagnoses reaching 96.7% (95% CI, 96.6%–96.9%) and 91.0% (95% CI, 90.9%–91.1%), respectively.

PPV declined significantly with increasing diagnosis position (**Figure 4**). Each step down was associated with a 0.12% decrease in PPV (95% CI, −0.14% to −0.11%; P < 0.001). This trend persisted after excluding principal diagnosis, though with a smaller effect size (β = −0.03%, 95% CI, −0.04% to −0.01%; P < 0.001).

**Figure 4.**
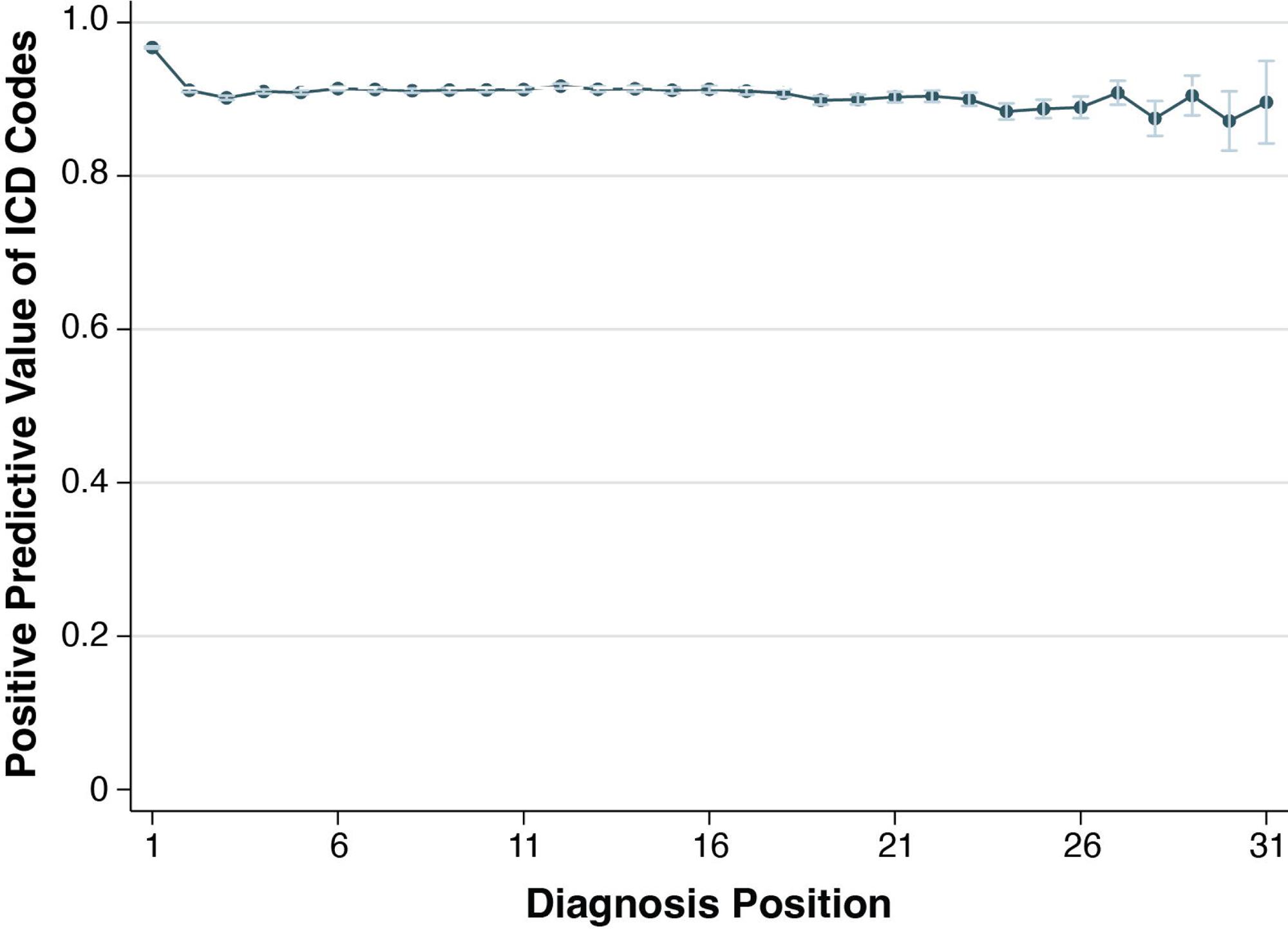
Positive Predictive Value of ICD-10 Codes by Diagnosis Position. The graph demonstrates the positive predictive value (PPV) of ICD-10 codes stratified by diagnosis position across 865,079 diagnoses from 64,764 hospital admissions. Each point represents the proportion of confirmed valid codes at that diagnosis position, with error bars indicating 95% confidence intervals. Diagnoses beyond position 31 (<0.1% of total) were excluded due to small sample sizes.

### Predictors of Inaccurate Real-World ICD-10-CM Assignments

We analyzed predictors of inaccurate ICD-10-CM assignments using the full dataset of 865,079 code assignments validated by the LLM system. This exploratory analysis aimed to identify clinical or administrative contexts where coding errors were more likely to occur, thereby informing opportunities for targeted quality improvement. To reduce bias from poorly documented codes (e.g., immunization, tobacco use), we restricted the analysis to the 68 most frequent codes (frequency >2,000) and excluded those in the lowest PPV quartile.

Surgical same-day admission was the strongest predictor of inaccuracy (adjusted OR = 2.10, 95% CI, 2.00–2.21), while hospice discharge was protective (adjusted OR = 0.75, 95% CI, 0.67–0.85). No demographic variables were significantly associated with inaccurate codes. Full results are presented in **Supplementary Figure S2**.

### Clinical Relevance of Secondary Diagnoses

Analysis of 611,841 correctly assigned ICD-10-CM secondary diagnoses revealed that only 32.1% (95% CI, 32.0%-32.2%) represented clinically active conditions requiring intervention during hospitalization, whether acute exacerbations of chronic conditions or new diagnoses. The remaining 67.9% captured baseline or historical conditions. Of the active diagnoses, 22.3% (95% CI, 22.1%–22.5%) were hospital-acquired, while 77.7% were present on admission. Full distribution details are provided in **Supplementary Table S3** and the online repository. This analysis reflects the second phase of the LLM audit workflow, in which the model was used separately to assess the clinical relevance of each diagnosis. This task provides data for researchers relying on secondary codes to identify comorbidities or hospital-acquired conditions.

### Resource Utilization

The LLM system demonstrated efficient resource use across validation tasks. For ICD code correctness, the mean per-chart cost was $0.097 (SD, $0.032) with an average processing time of 45.9 seconds (SD, 24.8). Mean input and output token lengths were 5,350 (SD, 1,642) and 1,887 (SD, 789), respectively. Evaluation of clinical relevance was more efficient, with a mean cost of $0.068 (SD, $0.023) and processing time of 19.0 seconds (SD, 12.8). Input length was similar (5,223 [SD, 1,632]), while output length was shorter (948 [SD, 441]).

## DISCUSSION

In this large-scale evaluation, we present the first comprehensive assessment of PPV for a broad range of diagnosis codes. Our findings not only offer a centralized dictionary of code-specific PPVs – an invaluable resource for investigators leveraging administrative datasets – but also reveal that, although ICD codes generally exhibit good accuracy, there is notable variability by condition type and code location (e.g., principal vs. secondary diagnoses). Furthermore, we demonstrate that a validated LLM system can serve as an exceptional tool for evaluating the validity of ICD codes, frequently matching or outperforming human reviewers. This suggests that LLM-enhanced auditing could substantially improve the reliability of ICD-coded data, thus strengthening both clinical research and administrative decision-making.

ICD-10-CM codes serve as the backbone of the majority of administrative healthcare databases, serving critical functions in clinical research, reimbursement, and policymaking.^15^ Despite their ubiquity, concerns regarding coding accuracy persist, with prior studies reporting widely divergent validity ranging from 53% to 98%.^12,13^ While various validation efforts have endeavored to establish code-specific reliabilities,^6,7^ no single study has comprehensively examined the accuracy of the ICD-10-CM codes in routine practice at scale. By evaluating 13,102 unique codes, our findings reveal a wide range of validities, providing a valuable resource for researchers and policymakers.

In assessing the clinical relevance of secondary diagnoses, our goal was not to evaluate their impact on treatment decisions – a standard coding criterion – but to determine whether they reflected active conditions (e.g., acute episodes or exacerbations) or historical/baseline health status. This distinction is critical for clinical research.^9^ For example, in studies of acute pancreatitis as a secondary diagnosis, distinguishing active episodes from historical mentions and identifying whether the condition was present on admission is essential. Overall, most secondary diagnoses (68%) were determined to be historical. However, when restricting the analysis to codes explicitly labeled as “acute,” nearly all (median, 100%; IQR, 95.5%-100%) were active conditions, suggesting that such codes can reliably be treated as “active” for research purposes. Certain codes indicative of acute illness, such as melena (93.1%) and hematemesis (96.9%), also demonstrated high rates of active disease. Most active conditions were present on admission, with only 7.1% (IQR, 0%–35.0%) emerging during hospitalization. These findings, including code-level PPV, offer a reference for researchers interpreting ICD-coded diagnoses.

While our primary focus was on validating LLM performance and summarizing overall code accuracy, future iterations of this system should include structured analysis of discordant cases – that is, disagreements between the LLM and human reviewers. Root cause analysis of such discrepancies could identify edge cases and systematic failure modes, providing valuable training signals for reinforcement learning with human feedback or other refinement techniques. This would allow for continuous improvement of the system and further enhance reliability in real-world deployment.

The study identified key risk factors for ICD-10-CM coding errors. Same-day surgical admissions were associated with increased risk. The lack of association with patient demographics or clinical complexity suggests workflow inefficiencies, rather than patient characteristics, as primary contributors. These findings support the need of automated auditing tools, such as LLM-based systems, to improve coding accuracy, particularly for high-risk admissions and secondary diagnoses.

Recent advancements in natural language processing have transformed the extraction of clinical features from unstructured data sources. Domain-specific models, trained on clinical datasets, have demonstrated their efficacy in clinical concept extraction.^16^ Similarly, large scale general-purpose LLMs have proven highly adept at identifying clinical events with remarkable accuracy.^17^ Although these technologies show immense promise, their application to automated validation of diagnostic coding systems remains underexplored. Far et al. recently illustrated this potential through reliable LLM-based classification of cirrhosis-related diagnoses,^18^ yet the broader implications for code auditing remain unaddressed. Our findings show that a rigorously structured LLM system not only outperforms traditional human review but also overcomes key challenges in large-scale administrative data validation. The cognitive burden of reviewing extensive documentation limits human consistency, whereas LLM-based auditing offers a scalable solution.

Although the LLM achieved accuracy comparable to or exceeding that of physician reviewers, the observed Cohen κ of 0.51 indicates only moderate agreement. This discrepancy likely reflects differences in decision patterns rather than inconsistent performance. Despite extensive prompt optimization, the LLM occasionally makes isolated factual or interpretive errors (“hallucinations”) but tends to demonstrate exceptional consistency and attention to detail—areas where human reviewers may vary more. Thus, the moderate κ reflects divergence in reasoning approaches rather than a shortcoming of either method.

Importantly, these findings support the use of LLMs as augmentative rather than replacement tools. LLM-based auditing can serve as a first-pass screening system to flag potential discrepancies, improve workflow efficiency, and enhance reproducibility, while physician oversight remains essential for contextual interpretation and adjudication of ambiguous or novel cases. Together, human–AI collaboration offers a balanced model for scalable, reliable ICD-10 validation.

Automated clinical coding remains a critical challenge in health informatics, complicated by the intrinsic high dimensionality of modern classification systems. While previous investigations have explored LLM applications for lower-dimensional tasks – such as Diagnosis-Related Group prediction – these approaches demonstrated limited real-world applicability, with error rates exceeding practical utility thresholds.^19^ In addition, Soroush et al. have reported that LLMs perform inadequately as medical coders: GPT-4 achieved 33.9% exact-match accuracy for ICD-10-CM codes when merely prompted with diagnosis descriptions.^20^ These shortcomings can be attributed to factors such as insufficient domain-specific ICD knowledge and the reliance on imperfect real-world data that may include erroneous ground truth labels.^21^ By curating high-quality, validated data, our work not only offers a more reliable benchmark but also provides robust training data that can improve LLM coding accuracy.

Currently, we focus on verifying whether assigned codes are correct (PPV) rather than identifying omitted diagnoses (NPV). While the LLM is adept at confirming assigned codes, generating new codes remains a next step. This validated dataset provides a strong foundation for developing LLMs capable of automated ICD-10-CM code generation.

Reproducibility and implementation across institutions present practical challenges. Our system was developed using GPT-4o within a secure institutional environment, and although we have provided the prompt template and model version, performance may vary with different LLM versions or institutional contexts. Prior to deployment elsewhere, local validation and prompt refinement are essential, as documentation patterns and coding errors can differ substantially across settings.

Access and privacy constraints may also limit the use of commercial LLMs in certain institutions or countries where data-sharing regulations prohibit cloud-based processing of protected health information. In such environments, alternative strategies could include deployment of locally hosted LLMs or the use of open-source models with comparable performance. While a performance gap historically existed between open and proprietary models, recent open-weight systems have demonstrated increasingly competitive capabilities. Another promising pathway involves fine-tuning smaller, secure LLMs on synthetic datasets generated by larger foundation models, thereby maintaining privacy while achieving task-specific optimization. These options provide practical pathways for adapting the described approach to diverse institutional and regulatory contexts, facilitating broader, secure adoption of LLM-based auditing frameworks.

This study’s key strengths include its unprecedented scale and rigorous methodology (a validated LLM with iterative prompt engineering and physician adjudication). The consistently high performance of the LLM-based system in validating ICD-10-CM codes shows a feasible, efficient way to enhance administrative data accuracy. Moreover, the granular code-level analysis and distinction between active and historical conditions enhance clinical relevance for researchers.

Several limitations warrant consideration. First, the MIMIC-IV database is derived from a single tertiary academic center, which may limit generalizability to other settings with different patient populations and coding practices. Coding practices can vary substantially across institutions, regions, and countries due to differences in documentation styles, billing priorities, coder training, and institutional policies. While our results provide a benchmark for real-world coding validity in a large academic hospital setting, further validation is needed in other care settings to determine the generalizability of our findings. Future studies applying LLM-based auditing tools across diverse datasets will be critical for assessing their broader applicability. Second, de-identification procedures could have removed important clinical details (e.g., Hashimoto’s thyroiditis). Third, although discharge summaries were available, other documents such as progress notes and procedural notes were not, potentially constraining the model’s ability to validate certain conditions. Fourth, a small amount of admissions with large numbers of diagnoses exceeded the LLM’s output token limit. Fifth, despite effort to address misalignment between LLM and human intentions through iterative prompt refinement, some degree of underfitting likely persists, as not all edge cases or error patterns can be preemptively captured in a prompt – a known limitation in the field of prompt-based model alignment. This residual error, along with intrinsic hallucination risk, contributed to the inaccurate predictions during the testing and deployment phases. Lastly, although the auditing system performed robustly overall, its higher sensitivity than specificity may slightly underestimate the validity of certain codes.

In summary, these findings highlight a broad, real-world perspective on ICD-10-CM validity and the potential of LLM-based auditing to improve coding reliability. By revealing variation in code performance, particularly for lower-position secondary diagnoses and same-day surgical admissions, this work outlines opportunities for targeted quality improvements. The LLM system’s consistency, scalability, and adaptability across diverse clinical contexts make it suitable for routine integration into both research and clinical workflows, ultimately enhancing research rigor, reimbursement accuracy, and patient care.

## AUTHORS CONTRIBUTIONS

Y.W.: Data collection, formal analysis, original draft writing; review and editing Y.S.: Formal analysis, original draft writing; R.S.: Data collection, formal analysis; I.N.: Data collection, formal analysis, methodology; Y.Y.: Data collection, analysis, writing – review and editing; T.S.: Data collection, formal analysis, methodology; Z.L.: Data collection, formal analysis; D.R.: Data collection, formal analysis; J.W.: Data collection, formal analysis; D.B.: Data collection, formal analysis; C.T.: Conceptualization; V.K.: Conceptualization, funding acquisition, supervision; Y.H.: Project administration, conceptualization, funding acquisition, supervision.

## ACKNOWLEDGMENTS

The authors thank the Mayo Clinic Cloud team for their infrastructure support and all contributors to the MIMIC-IV database.

## DATA AVAILABILITY

Aggregated results are provided in the manuscript, Supplementary Materials, and the online repository cited in the text. De-identified, line-level validation outputs (LLM judgments and adjudication labels) are available from the corresponding author upon reasonable request and will be shared pending Mayo Clinic approval and a data use agreement. Source EHR data derive from MIMIC-IV; access requires submitting a data use application and completing the required training with the MIMIC team.

## REFERENCES

1. World Health Organization. WHO Mortality Database: Tables, GD. Accessed March 2, 2023. https://www.who.int/data/data-collection-tools/who-mortality-database.

2. World Health Organization. International Statistical Classification of Diseases and Related Health Problems: Tenth Revision. 2nd Ed. Geneva: World Health Organization; 2004. .

3. Centers for Medicare & Medicaid Services. National Health Expenditures 2022 Highlights. Published 2023. Accessed February 8, 2024. https://www.cms.gov/files/document/highlights.pdf.

4. Centers for Medicare & Medicaid Services. 2022 Medicare Fee-for-Service Supplemental Improper Payment Data. Published December 8, 2022. Accessed February 8, 2024. https://www.cms.gov/data-research/monitoring-programs/improper-payment-measurement-programs/comprehensive-error-rate-testing-cert/cert-reports/2022-medicare-fee-service-supplemental-improper-payment-data.

5. Agency for Healthcare Research and Quality. Healthcare Cost and Utilization Project (HCUP). Rockville, MD. Accessed June 3, 2025. https://www.hcup-us.ahrq.gov.

6. Columbo JA, Daya N, Colantonio LD, et al. Derivation and Validation of ICD-10 Codes for Identifying Incident Stroke. JAMA Neurol. 2024;81(8):875–881. doi:10.1001/jamaneurol.2024.2044

7. Antoon JW, Stopczynski T, Amarin JZ, et al. Accuracy of Influenza ICD-10 Diagnosis Codes in Identifying Influenza Illness in Children. JAMA Netw Open. 2024;7(4):e248255. doi:10.1001/jamanetworkopen.2024.8255

8. Khokhar B, Jette N, Metcalfe A, et al. Systematic review of validated case definitions for diabetes in ICD-9-coded and ICD-10-coded data in adult populations. BMJ Open. 2016;6(8):e009952. doi:10.1136/bmjopen-2015-009952

9. Khera R, Angraal S, Couch T, et al. Adherence to Methodological Standards in Research Using the National Inpatient Sample. JAMA. 2017;318(20):2011–2018. doi:10.1001/jama.2017.17653

10. Omiye JA, Gui H, Rezaei SJ, Zou J, Daneshjou R. Large Language Models in Medicine: The Potentials and Pitfalls□: A Narrative Review. Ann Intern Med. 2024;177(2):210–220. doi:10.7326/M23-2772

11. OpenAI. GPT-4 Technical Report. Accessed February 3, 2025. https://openai.com/research/gpt-4. .

12. Dong H, Falis M, Whiteley W, et al. Automated clinical coding: what, why, and where we are? NPJ Digit Med. 2022;5(1):159. doi:10.1038/s41746-022-00705-7

13. Burns EM, Rigby E, Mamidanna R, et al. Systematic review of discharge coding accuracy. J Public Health (Oxf). 2012;34(1):138–148. doi:10.1093/pubmed/fdr054

14. Johnson AEW, Bulgarelli L, Shen L, et al. MIMIC-IV, a freely accessible electronic health record dataset. Sci Data. 2023;10(1):1. doi:10.1038/s41597-022-01899-x

15. Quan H, Sundararajan V, Halfon P, et al. Coding algorithms for defining comorbidities in ICD-9-CM and ICD-10 administrative data. Med Care. 2005;43(11):1130–1139. doi:10.1097/01.mlr.0000182534.19832.83

16. Yang X, Chen A, PourNejatian N, et al. A large language model for electronic health records. NPJ Digit Med. 2022;5(1):194. doi:10.1038/s41746-022-00742-2

17. Wang Y, Huang Y, Nimma IR, et al. Validation of GPT-4 for clinical event classification: A comparative analysis with ICD codes and human reviewers. J Gastroenterol Hepatol. 2024;39(8):1535–1543. doi:10.1111/jgh.16561

18. Far AT, Bastani A, Lee A, et al. Evaluating the positive predictive value of code-based identification of cirrhosis and its complications utilizing GPT-4. Hepatology. Published online October 8, 2024. doi:10.1097/HEP.0000000000001115

19. Wang H, Gao C, Dantona C, Hull B, Sun J. DRG-LLaMA□: tuning LLaMA model to predict diagnosis-related group for hospitalized patients. NPJ Digit Med. 2024;7(1):16. doi:10.1038/s41746-023-00989-3

20. Soroush A, Glicksberg BS, Zimlichman E, et al. Large Language Models Are Poor Medical Coders — Benchmarking of Medical Code Querying. NEJM AI. 2024;1(5). doi:10.1056/AIdbp2300040

21. Kwan K. Large language models are good medical coders, if provided with tools. Published online July 6, 2024.

