## Supplemental Material for "Validation of 13,102 ICD-10-CM Codes Using a Large Language Model-Based System"

***LLM System Implementation Details***

GPT-4o-2024-05-13 was the deployed model version during our analysis period from November 1, 2024, to December 31, 2024 on Mayo Clinic Cloud. Our large language model (LLM) system implementation incorporated technical specifications designed to ensure consistent and reliable code validation. The system utilized asynchronous (application programming interface) API calls with a concurrency limit of 54 simultaneous requests to maintain system stability while optimizing throughput. Each API request was configured with a temperature setting of 0.7 to balance variability (necessary for majority voting) and consistency in the LLM's reasoning process. The LLM provider imposed a maximum output token limit of 4,096 and a context window limit of 128,000 tokens at the time of analysis. To ensure reliable processing of large medical records, we implemented message size monitoring and automatic handling of API rate limits.

Our prompt engineering process focused on key elements that significantly impact analysis quality. The system prompt established the LLM's role as an experienced attending physician with 30 years of clinical practice, emphasizing expertise in diagnostic criteria and common documentation pitfalls. We implemented specific guidance for handling diagnostic ambiguity, including rules for accepting conceptually equivalent codes and managing unspecified conditions. The prompt included detailed instructions for analyzing documentation support, considering diagnostic criteria from specialty societies, and evaluating common medical student mistakes in diagnosis—factors crucial for accurate code validation.

While we aimed for high coding accuracy, our approach was intentionally slightly more flexible than strict official guidelines. The primary objective was to assess whether the International Classification of Diseases (ICD) codes meaningfully correlated with the clinical narrative. Accordingly, we accepted codes labeled as “other” or “unspecified” when more specific information was present in the chart, provided the documentation supported the general category. We also assumed that a diagnosis referred to its most common type unless otherwise specified. Additionally, we considered codes to be valid even when not explicitly documented, as long as they could be reasonably inferred from the available clinical data.

The output format was strictly controlled through a structured JavaScript Object Notation (JSON) template that required explicit documentation of thought processes and reasoning for each code evaluation. This structured approach ensured comprehensive coverage of key diagnostic considerations while maintaining consistency across evaluations. The system incorporated cost calculation based on token usage, with rates of 0.0025 per prompt token and 0.01 per completion token, enabling monitoring of computational resource utilization.

To optimize response processing, we implemented robust error handling and response validation protocols. These included automated checks for missing fields, JSON syntax validation, and standardized processing of API responses. This comprehensive approach to system implementation ensured reliable and consistent code validation across large-scale medical record analysis.

The complete system prompt template and implementation details are available upon reasonable request to the corresponding author.

***Data Availability***

Validation results for individual ICD-10-CM codes—including overall positive predictive value (PPV), PPV for codes reflecting an active issue (i.e., deviating from baseline health status), and PPV for active issues occurring after admission—are available at the following online repository: https://huggingface.co/datasets/wangyichen25/ICD-10-CM_Validation_Results/viewer. Line-level clinical data can be accessed by submitting application through the MIMIC-IV database at https://physionet.org/content/mimiciv/3.1/.

***Input Example***

Below is an example of input provided to a large language model (LLM). The raw Markdown format is shown intentionally. Text enclosed in triple quotation marks represents the message sent to the LLM. Content within curly brackets (“{}”) provides explanatory notes for readers of this paper and is not part of the actual input sent to the LLM.

The following is the system message, which sets the behavior and role of the LLM throughout the interaction.

"""

You are a loyal assistant who follows my instruction closely. You are an academic attending physician who have practiced for 30 years and has extensive experience in clinical diagnosis and research. You know clearly about diagnostic criteria for all medical conditions. You have rich educational experience and knows common mistakes medical students tend to make. You have detailed knowledge about ICD-10 codes. You are helping me to audit if an ICD-10 code is assigned correctly for clinical research.
"""

The following is the user message, which represents the input or query provided by the user to prompt a response from the LLM.

"""

Please review the medical record below carefully and assess the accuracy of the assigned ICD-10-CM codes. Your precise evaluation is essential, as accurate diagnoses are critical for research, guiding public policy, and improving patient care. Only mark a code as incorrect if you have a clear and well-founded reason, as these codes are assigned by expert coders.

---

**Demographics:**

Race: {race}

Gender: {gender}

Age: {age}

**Medical Record:**

{raw discharge summary, which often includes pertinent radiology reports, labs, medications}

**Additional Clinical Information:**

{Additional clinical information extracted from structured data are placed here. Include: body mass index, laboratory test results, and vital signs [heart rate, blood pressure (systolic, diastolic, and mean arterial pressure), respiratory rate, oxygen saturation, body temperature, and supplemental oxygen flow rate]. The dataset encompassed the complete range of recorded minimum and maximum values for laboratory tests and vital signs during hospitalization. Microbiology culture results, including antibiotic susceptibility interpretations, were also included. Furthermore, additional clinical parameters, such as fluid balance calculations, urine output, and device-related metrics (e.g., ventilator settings, central venous pressure), were provided if available.}

---

**Assigned Codes for Review:**

{Existing real-world coder-assigned ICD-10 codes.

Example:

L02413 [dx1] - Cutaneous abscess of right upper limb

F1120 [dx2] - Opioid dependence, uncomplicated

F17210 [dx3] - Nicotine dependence, cigarettes, uncomplicated}

---

**Instructions:**

**Understand Code Format:**

- Codes are formatted as: [ICD code number] [sequence] - [ICD code description], where [sequence] indicates diagnosis order ([dx1], [dx2], etc.; [dx1] is the principal diagnosis).

- Focus on the description, not the code number.

**Analysis Steps:**

Before analyzing the codes, in the "thought" section, outline your approach to auditing the ICD-10 codes based on the specific medical record provided.

For each code:

1. Consider the diagnosing criteria based on clinical guidelines from major specialty societies.

2. Reflect common mistakes medical students tend to make in making the diagnosis (e.g., hypertension from one elevated BP reading, UTI from UA without documentation of symptoms and diagnosis by the provider).

3. Extract relevant documentation explicitly authored by the provider.

4. Confirm that the diagnosis is clearly documented and clinically supported, or identify any gaps or contradictions.

5. If the code is incorrect but the condition exists and is documented, provide the correct alternative code that accurately reflects the documented condition.

**Code Analysis Guidelines:**

1. **Accept Approximations:**

- Recognize that ICD-10 may not have exact matches for all medical terms; accept conceptually equivalent codes even if wording differs. Examples:

- Use the best available code for a condition, even with minor anatomical distinctions.

- "Acute kidney injury (AKI)" == "Acute kidney failure"

- "Decompensated cirrhosis" == "Liver failure"

- "Coronary artery disease" ≈ "Atherosclerotic heart disease of native coronary artery"

- "Encephalopathy" ≈ "Altered mental status/confusion"

2. **Unspecified Conditions:**

- Using "other" or "unspecified" conditions is acceptable if the condition exists. For example, "Other postprocedural cardiac functional disturbances following cardiac surgery" for postoperative atrial fibrillation.

3. **Assuming Common Diagnoses:**

- It is acceptable to assume a diagnosis refers to its most common type unless specified otherwise, provided this assumption does not significantly impact patient care.

Examples: arthritis → osteoarthritis; hypertension → essential hypertension; diabetes → type 2 diabetes mellitus; stroke → cerebral infarct...

4. **Include Resolved/Improved Conditions:**

- Code conditions treated during admission, even if resolved by discharge (e.g., resolved acute kidney injury).

5. **Combination Diagnoses:**

- Accept combination codes (e.g., "hypertension with heart failure" or "hypertensive CKD") if all listed conditions are present. **Assume causal relationships by default** unless explicitly stated otherwise by the provider.

6. **Documented Diagnoses:**

- Conditions documented by the provider are ALWAYS acceptable to code.

- Coding conditions from the patient's past medical history as secondary diagnoses is generally acceptable.

7. **Probable, Suspected, or Likely Diagnoses:**

- Code conditions documented as "probable," "suspected," or "likely" as if they are confirmed.

8. **Undocumented Diagnoses:**

- If a code lacks clear documentation, assess whether it can be reasonably inferred from the clinical data. Refer to established clinical guidelines to ensure the inference aligns with evidence-based standards. Be cautious of common diagnostic pitfalls medical students might encounter, such as relying on isolated findings or testing results without incorporating clinical context.

9. **Provide Correct Alternatives for Incorrect Codes:**

- If a code is incorrect, provide the correct alternative code unless there is no basis in the chart.

\### **Output Format:**

{

"thought": "[thought process]",

"dx1_reasoning": "1. [relevant documentation]; 2. [The diagnosing criteria for [ICD-10 code description] is...]; 3. [Common mistakes medical students make in diagnosing [ICD-10 code description] include [pitfalls]; there is/is no such mistake here]; 4. [There is sufficient evidence/contraindication based on the chart review for coding [ICD-10 code description], including... Therefore, [ICD-10 code description] is a correct/incorrect code here]; 5. [(ONLY IF THE CODE IS INCORRECT) The correct ICD-10-CM code should be...]",

"dx1_correct": 1, // Use 1 if the code is correct, 0 if incorrect

[continue for the rest of the diagnostic codes]

}

**Key Notes:**

- Do not reject a correct code solely because an exact match is absent; an exact match may not exist in ICD-10.

- Strictly follow the output JSON format. DO NOT USE MARKDOWN. Do not output anything other than the JSON.

- Do not nitpick.

- Use clear and concise language.

"""

***LLM Response Example***

{

"thought": "I will review the medical record in detail, focusing on the documented diagnoses and clinical findings. For each ICD-10 code, I will consider the diagnosing criteria based on clinical guidelines, reflect on common diagnostic mistakes, extract relevant documentation from the record, and ensure that the diagnosis is clearly documented and clinically supported. If a code is incorrect but the condition is documented, I will provide the correct alternative code.",

"dx1_reasoning": "1. Relevant documentation: The patient reports having melena for the past 2 months, and this is confirmed during the review of systems. 2. The diagnosing criteria for melena include the presence of black, tarry stools indicative of gastrointestinal bleeding. 3. Common mistakes medical students make in diagnosing melena include mistaking other causes of black stools (such as iron supplements or certain foods) for melena, but there is no such mistake here. 4. There is sufficient evidence based on the chart review for coding melena, including the patient's report and clinical context of GI bleeding. Therefore, melena is a correct code here.",

"dx1_correct": 1,

"dx2_reasoning": "1. Relevant documentation: The patient was found to have a hemoglobin level of 5.4 and received multiple transfusions for anemia due to gastrointestinal bleeding. 2. The diagnosing criteria for acute posthemorrhagic anemia include a significant drop in hemoglobin/hematocrit levels due to acute blood loss. 3. Common mistakes medical students make in diagnosing acute posthemorrhagic anemia include not distinguishing it from chronic anemia, but this is clearly an acute case. 4. There is sufficient evidence based on the chart review for coding acute posthemorrhagic anemia, including the documented hemoglobin level and transfusions. Therefore, acute posthemorrhagic anemia is a correct code here.",

"dx2_correct": 1,

"dx3_reasoning": "1. Relevant documentation: The patient has a history of asthma and is continued on albuterol inhaler. 2. The diagnosing criteria for unspecified asthma, uncomplicated include a documented history of asthma without current exacerbation or complications. 3. Common mistakes medical students make in diagnosing asthma include not specifying the type or severity, but this is an unspecified case without complications. 4. There is sufficient evidence based on the chart review for coding unspecified asthma, uncomplicated, including the patient's history and medication. Therefore, unspecified asthma, uncomplicated is a correct code here.",

"dx3_correct": 1,

"dx4_reasoning": "1. Relevant documentation: The patient has a large hiatal hernia confirmed by multiple imaging studies. 2. The diagnosing criteria for diaphragmatic hernia without obstruction or gangrene include the presence of a hernia in the diaphragm, confirmed by imaging, without signs of obstruction or gangrene. 3. Common mistakes medical students make in diagnosing diaphragmatic hernia include not distinguishing it from other types of hernias, but this case is specific to the diaphragm. 4. There is sufficient evidence based on the chart review for coding diaphragmatic hernia without obstruction or gangrene, including the documented imaging findings. Therefore, diaphragmatic hernia without obstruction or gangrene is a correct code here.",

"dx4_correct": 1,

"dx5_reasoning": "1. Relevant documentation: There is no mention of an immunization encounter during this hospitalization. 2. The diagnosing criteria for encounter for immunization require documentation of the patient receiving or being evaluated for immunization. 3. Common mistakes medical students make in diagnosing encounter for immunization include assuming it based on routine care without specific documentation. 4. There is no evidence based on the chart review for coding encounter for immunization. Therefore, encounter for immunization is an incorrect code here. 5. There is no alternative code needed as this condition was not documented.",

"dx5_correct": 0

}

***Input Example – Active vs. Inactive Diagnoses***

Below is an example of input provided to a large language model (LLM) for active vs. inactive diagnoses. The raw Markdown format is shown intentionally. Text enclosed in triple quotation marks represents the message sent to the LLM. Content within curly brackets (“{}”) provides explanatory notes for readers of this paper and is not part of the actual input sent to the LLM.

The following is the system message, which sets the behavior and role of the LLM throughout the interaction.

"""

You are a loyal assistant who follows my instruction closely. You are an academic attending physician who have practiced for 30 years and has extensive experience in clinical diagnosis and research. You know clearly about diagnostic criteria for all medical conditions. You have riched educational experience and knows common mistakes medical students tend to make. You have detailed knowledge about ICD-10 codes. You are helping me to audit if an ICD-10 code represents an active issue for a clinical research.

"""

The following is the user message, which represents the input or query provided by the user to prompt a response from the LLM.

"""

You are provided with a set of ICD-10-CM diagnosis codes that have been validated as correctly documented in the patient's chart. Your goal is to classify each condition into one of the categories based on its activeness and timing relative to the admission:

1. Activeness: Determine whether the condition is "active" or "not active".

- Active (Code "1")

- Not Active (Code "0")

2. Timing (only if the condition is active):

- Active conditions that were present at the time of admission: Assign "0" for "Active on Admission."

- Active that developed or were first recognized after admission: Assign "1" for "Active after Admission."

Definitions and Guidance:

- "Active conditions" encompasses medical conditions that represent a significant change from a patient's baseline health status. These can be:

• Acute (sudden onset)

• Subacute (developing over days to weeks)

• Acute on chronic (sudden worsening of an existing condition)

• Subacute on chronic (gradual worsening of an existing condition)

Any condition requiring urgent medical intervention or close monitoring is considered active. However, not all active conditions necessarily require intervention. Here are some examples:

• Myocardial infarction (heart attack) that has just occurred

• Stroke with new onset of symptoms

• Recently diagnosed pneumonia requiring treatment

• New traumatic injuries from an accident

• COPD exacerbation causing marked decline in breathing function

• Rheumatoid arthritis flare with noticeable increase in joint pain and swelling beyond usual symptoms

• Sudden deterioration of kidney function in a patient with existing kidney disease

- "Not active" (or stable chronic) conditions are those that are long-standing and currently stable, with no acute decompensation or exacerbation. These conditions are managed at their baseline level. Examples include:

• Stable, well-controlled hypertension

• Longstanding, stable type 2 diabetes mellitus without acute complications

• Chronic kidney disease stable at baseline creatinine level

• Old myocardial infarction with no recent changes in cardiac status

Because these are actual hospital admissions, be especially careful before deciding that a principal diagnosis (the main reason for admission) is "0" (not active or not active on admission). Any conclusion that the principal diagnosis did not represent an active illness on admission should be strongly justified with a clear clinical rationale.

"Chronic" in code description ≠ automatically inactive; "Acute" in code description ≠ automatically active. Base classification on clinical documentation, not code terminology

Why This Matters:

- Researchers aiming to identify in-hospital events can use this analysis to exclude diagnoses that are merely historical carry-overs from previous visits.

- This ensures that conditions counted are actually active and relevant, helping researchers produce more accurate data on incidence rates (e.g., true acute MI rates), resource utilization, and outcomes.

- Policymakers and quality improvement teams may rely on these purified data sets to better understand which conditions are active during hospitalization and to optimize healthcare resource allocation.

- Healthcare analysts can study patterns of care, length-of-stay, and costs associated specifically with active conditions, without inflated numbers due to historical diagnoses that no longer reflect current acute needs.

---

**Demographics:**

Race: {race}

Gender: {gender}

Age: {age}

**Medical Record:**

{raw discharge summary, which often includes pertinent radiology reports, labs, medications}

**Additional Clinical Information:**

{Additional clinical information extracted from structured data are placed here. Include: body mass index, laboratory test results, and vital signs [heart rate, blood pressure (systolic, diastolic, and mean arterial pressure), respiratory rate, oxygen saturation, body temperature, and supplemental oxygen flow rate]. The dataset encompassed the complete range of recorded minimum and maximum values for laboratory tests and vital signs during hospitalization. Microbiology culture results, including antibiotic susceptibility interpretations, were also included. Furthermore, additional clinical parameters, such as fluid balance calculations, urine output, and device-related metrics (e.g., ventilator settings, central venous pressure), were provided if available.}

---

**Assigned Codes for Review:**

{Existing real-world coder-assigned ICD-10 codes.

Example:

L02413 [dx1] - Cutaneous abscess of right upper limb

F1120 [dx2] - Opioid dependence, uncomplicated

F17210 [dx3] - Nicotine dependence, cigarettes, uncomplicated}

---

For each code, provide a brief reasoning process, then assign the appropriate numeric code based on the definitions above.

You must produce a JSON object **WIHTOUT MARKDOWN** with the following fields:

{

"thought": "[Describe your overall reasoning process briefly]",

"dxN_reasoning": "1. [Relevent documentation]; 2. [Why this condition is active or not active]; 3. [If this condition is active, did it present on admission or after admission]; 4. [Final code assignment]",

"dxN_is_active": 0 or 1, // 0 = Not active, 1 = Active

"dxN_is_active_after_admission": 0 or 1, // 0 if 'is_active' is 0 or if the active condition exist on admission; 1 if the active condition developed after admission

[continue for the rest of the diagnostic codes]

}

Focus solely on the activeness (or lack thereof) and timing of the conditions. The diagnoses have already been validated as correct, so do not question their existence or accuracy. If you determine a condition is not active, ensure that you have a solid justification, especially for the principal diagnosis.

Return only the JSON object as specified, with no extra commentary.

YOU ARE NOT TO USE MARKDOWN. DO NOT USE MARKDOWN SYNTAX IN THE RESPONSE.

Attention: The sequence of provided diagnostic codes (dxN) may not be continuous and may not start from dx1. Ensure your output aligns accurately with the sequence provided.

"""

**Supplementary Figure S1.** Distribution of Positive Predictive Values for ICD-10-CM Codes in Clinical Documentation. Box plot showing the distribution of positive predictive values (PPV) for ICD-10 diagnosis codes in clinical documentation. The horizontal line within the box represents the median PPV. The lower and upper edges of the box represent the first (25th percentile) and third (75th percentile) quartiles, respectively. The whiskers extend to the most extreme data points within 1.5 times the interquartile range. Points beyond the whiskers represent individual outliers.
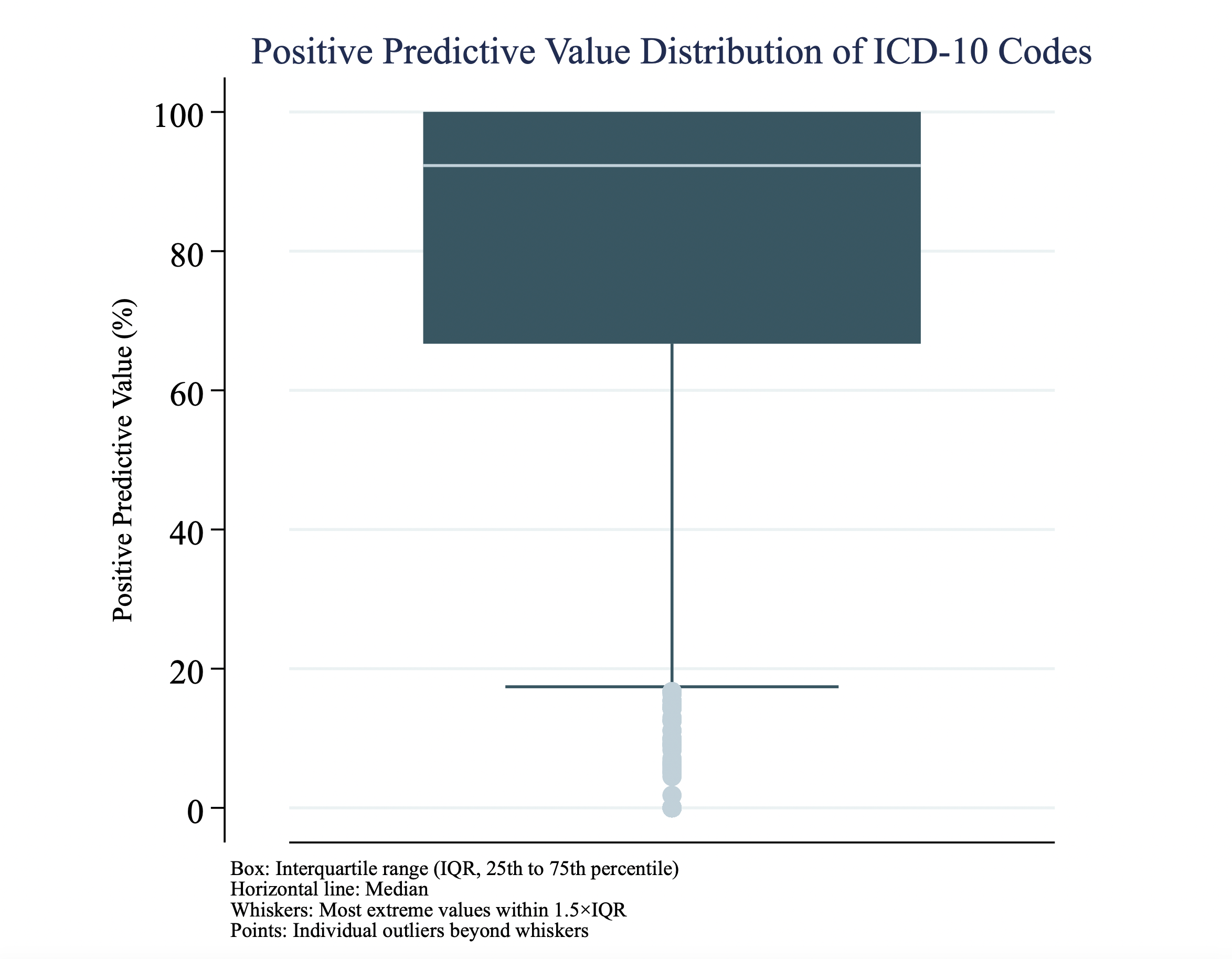

**Supplementary Figure S2**. Adjusted Odds Ratios for Predictors of Incorrect ICD-10-CM Assignment. Forest plot showing odds ratios (ORs) and 95% confidence intervals (CIs) from multivariable logistic regression analysis. Variables are grouped by clinical/administrative class and sorted by ascending OR within each class. The regression model adjusts for ICD code distribution using the 68 most frequently occurring codes (frequency >2000), although these adjustments are not displayed in the forest plot. Reference categories (gray squares), which are most frequent in each class, include: White race, direct admission location, elective admission type, home discharge location, English language, and private insurance. Odds ratios are presented on a log scale. Asterisks (*) denote statistically significant associations (p < 0.05).

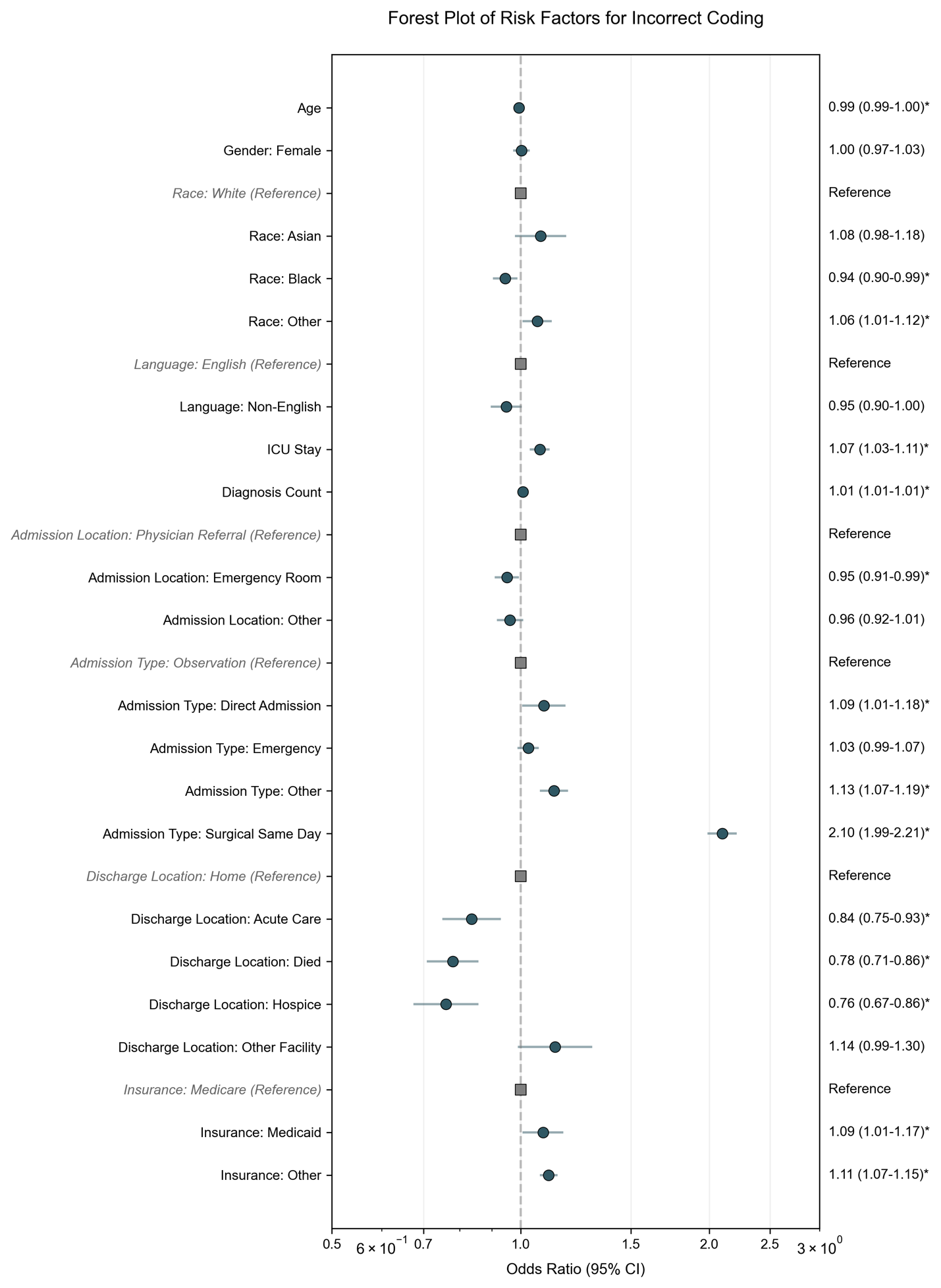

| **Supplementary Table S1.** Diagnostic Performance Metrics Stratified by ICD Code Frequency, Comparison Between GPT and Human Reviewers | | | | |
| --- | --- | --- | --- | --- |
| Performance Metric | Reviewer | Overall (N=971) | Top 10% Frequent (N=429) | Bottom 10% Frequent (N=386) |
| Sensitivity | GPT | 98.5% (97.4-99.2) | 97.7% (95.5-99.0) | 99.1% (97.3-99.8) |
|  | Human | 98.3% (97.1-99.0) | 98.0% (95.8-99.2) | 98.8% (96.9-99.7) |
| Specificity | GPT | 94.5% (89.8-97.4) | 95.2% (88.1-98.7) | 93.4% (84.1-98.2) |
|  | Human | 91.6% (86.3-95.3) | 93.0% (85.4-97.4) | 88.9% (78.4-95.4) |
| PPV | GPT | 98.9% (97.9-99.5) | 98.8% (97.0-99.7) | 98.8% (96.9-99.7) |
|  | Human | 98.3% (97.1-99.0) | 98.2% (96.2-99.4) | 97.9% (95.6-99.1) |
| NPV | GPT | 92.8% (87.7-96.2) | 90.8% (82.7-95.9) | 95.0% (86.1-99.0) |
|  | Human | 91.6% (86.3-95.3) | 92.0% (84.1-96.7) | 93.3% (83.8-98.2) |
| ROC Area | GPT | 0.96 (0.95-0.98) | 0.96 (0.94-0.99) | 0.96 (0.93-0.99) |
|  | Human | 0.95 (0.93-0.97) | 0.95 (0.93-0.98) | 0.94 (0.90-0.98) |
| Methods: This analysis stratified ICD codes based on their frequency of occurrence in the dataset. The top 10% frequent codes represent those appearing most often, while the bottom 10% represent the least frequent codes. Ground truth was used as the reference standard for all comparisons. Values are presented as point estimates with 95% confidence intervals in parentheses. PPV = positive predictive value; NPV = negative predictive value; ROC = receiver operating characteristic. | | | | |

| **Supplementary Table S2.** Analysis of Potential Predictors for Incorrect LLM System Judgments in ICD-10 Code Validation | | |
| --- | --- | --- |
| Characteristic | Odds Ratio (95% CI) | P-value |
| Patient Demographics |  |  |
| Age (per year) | 1.00 (0.97-1.03) | 0.925 |
| Female sex | 0.43 (0.14-1.29) | 0.132 |
| Race/Ethnicity* |  |  |
| - White | Reference | - |
| - Black | 1.26 (0.36-4.37) | 0.717 |
| - Other | 0.44 (0.06-3.36) | 0.43 |
| Language |  |  |
| - English | Reference | - |
| - Non-English | 1.20 (0.35-4.13) | 0.775 |
| Clinical Characteristics |  |  |
| Number of diagnoses | 1.06 (0.99-1.13) | 0.093 |
| Length of stay (days) | 1.01 (0.95-1.07) | 0.761 |
| ICU admission | 1.02 (0.37-2.83) | 0.962 |
| ICU length of stay† | 1.13 (0.95-1.34) | 0.167 |
| Admission Characteristics | |  |
| Admission Type |  |  |
| - Observation | Reference | - |
| - Emergency via ED | 1.87 (0.67-5.21) | 0.234 |
| - Direct admission | 0.60 (0.07-4.87) | 0.634 |
| - Surgical same day | 1.68 (0.44-6.46) | 0.447 |
| - Other | 1.85 (0.38-8.92) | 0.442 |
| Admission Source |  |  |
| - Physician referral | Reference | - |
| - Emergency room | 2.12 (0.75-6.04) | 0.158 |
| - Transfer | 1.30 (0.32-5.27) | 0.713 |
| - Other | 0.96 (0.24-3.89) | 0.958 |
| Insurance Status‡ |  |  |
| - Medicare | Reference | - |
| - Other | 1.12 (0.47-2.66) | 0.798 |
| *Asian and Hispanic categories were omitted due to perfect prediction. †Analysis limited to ICU patients (n=227). ‡Medicaid category was omitted due to perfect prediction. Note: All analyses were conducted using logistic regression with incorrect LLM judgment as the dependent variable. Each characteristic was analyzed in a univariate model due to the low event rate. | | |

| **Supplemental Table S3.** Clinical Activity Analysis of Top 50 ICD-10-CM Diagnoses | | | | | | |
| --- | --- | --- | --- | --- | --- | --- |
| ICD-10-CM Code | Code Description | Total Cases | Active Cases | Active Rate (%) | Post-Admission Active Cases | Post-Admission Rate (%) |
| I10 | Essential (primary) hypertension | 20730 | 1044 | 5 | 41 | 3.9 |
| E785 | Hyperlipidemia, unspecified | 19689 | 19 | 0.1 | 4 | 21.1 |
| K219 | Gastro-esophageal reflux disease without esophagitis | 12924 | 226 | 1.7 | 21 | 9.3 |
| I2510 | Atherosclerotic heart disease of native coronary artery without angina pectoris | 9151 | 230 | 2.5 | 4 | 1.7 |
| F329 | Major depressive disorder, single episode, unspecified | 9078 | 556 | 6.1 | 17 | 3.1 |
| F419 | Anxiety disorder, unspecified | 7966 | 1009 | 12.7 | 49 | 4.9 |
| Z87891 | Personal history of nicotine dependence | 7654 | 0 | 0 | 0 | N/A |
| E039 | Hypothyroidism, unspecified | 6909 | 113 | 1.6 | 18 | 15.9 |
| Z7901 | Long term (current) use of anticoagulants | 6811 | 8 | 0.1 | 1 | 12.5 |
| Z794 | Long term (current) use of insulin | 6445 | 12 | 0.2 | 2 | 16.7 |
| E119 | Type 2 diabetes mellitus without complications | 6271 | 114 | 1.8 | 32 | 28.1 |
| N179 | acute kidney failure/injury/injury, unspecified | 5965 | 5870 | 98.4 | 1779 | 30.3 |
| D649 | Anemia, unspecified | 5270 | 2088 | 39.6 | 183 | 8.8 |
| E669 | Obesity, unspecified | 5130 | 3 | 0.1 | 0 | 0 |
| I4891 | Unspecified atrial fibrillation | 5027 | 1850 | 36.8 | 854 | 46.2 |
| Z7902 | Long term (current) use of antithrombotics/antiplatelets | 4803 | 1 | 0 | 0 | 0 |
| G4733 | Obstructive sleep apnea (adult) (pediatric) | 4655 | 41 | 0.9 | 14 | 34.1 |
| J45909 | Unspecified asthma, uncomplicated | 4510 | 139 | 3.1 | 13 | 9.4 |
| J449 | Chronic obstructive pulmonary disease, unspecified | 3920 | 331 | 8.4 | 17 | 5.1 |
| Z66 | Do not resuscitate | 3782 | 10 | 0.3 | 6 | 60 |
| E1122 | Type 2 diabetes mellitus with diabetic chronic kidney disease | 3520 | 111 | 3.2 | 4 | 3.6 |
| N189 | Chronic kidney disease, unspecified | 3505 | 40 | 1.1 | 0 | 0 |
| I129 | Hypertensive chronic kidney disease with stage 1 through stage 4 chronic kidney disease, or unspecified chronic kidney disease | 3455 | 42 | 1.2 | 2 | 4.8 |
| D62 | Acute posthemorrhagic anemia | 3433 | 3422 | 99.7 | 1749 | 51.1 |
| Z955 | Presence of coronary angioplasty implant and graft | 3366 | 0 | 0 | 0 | N/A |
| I252 | Old myocardial infarction | 3353 | 0 | 0 | 0 | N/A |
| Y929 | Unspecified place or not applicable | 3232 | 3 | 0.1 | 0 | 0 |
| G8929 | Other chronic pain | 3229 | 166 | 5.1 | 1 | 0.6 |
| E871 | Hypo-osmolality and hyponatremia | 3180 | 2858 | 89.9 | 416 | 14.6 |
| Z86718 | Personal history of other venous thrombosis and embolism | 3127 | 0 | 0 | 0 | N/A |
| Z8673 | Personal history of transient ischemic attack (TIA), and cerebral infarction without residual deficits | 3104 | 0 | 0 | 0 | N/A |
| F17210 | Nicotine dependence, cigarettes, uncomplicated | 3050 | 55 | 1.8 | 0 | 0 |
| N390 | Urinary tract infection, site not specified | 3022 | 2935 | 97.1 | 1146 | 39 |
| N400 | Benign prostatic hyperplasia without lower urinary tract symptoms | 2955 | 5 | 0.2 | 1 | 20 |
| K5900 | Constipation, unspecified | 2823 | 1217 | 43.1 | 204 | 16.8 |
| N183 | Chronic kidney disease, stage 3 (moderate) | 2764 | 21 | 0.8 | 1 | 4.8 |
| Z951 | Presence of aortocoronary bypass graft | 2632 | 0 | 0 | 0 | N/A |
| M109 | Gout, unspecified | 2578 | 171 | 6.6 | 85 | 49.7 |
| G4700 | Insomnia, unspecified | 2543 | 312 | 12.3 | 19 | 6.1 |
| I480 | Paroxysmal atrial fibrillation | 2511 | 981 | 39.1 | 331 | 33.7 |
| E872 | Acidosis | 2501 | 2388 | 95.5 | 351 | 14.7 |
| D696 | Thrombocytopenia, unspecified | 2271 | 1334 | 58.7 | 244 | 18.3 |
| D72829 | Elevated white blood cell count, unspecified | 2225 | 1853 | 83.3 | 350 | 18.9 |
| I5032 | Chronic diastolic (congestive) heart failure | 2204 | 118 | 5.4 | 1 | 0.8 |
| Z86711 | Personal history of pulmonary embolism | 2167 | 0 | 0 | 0 | N/A |
| M810 | Age-related osteoporosis without current pathological fracture | 2061 | 0 | 0 | 0 | N/A |
| D509 | Iron deficiency anemia, unspecified | 2024 | 720 | 35.6 | 20 | 2.8 |
| I110 | Hypertensive heart disease with heart failure | 2003 | 434 | 21.7 | 1 | 0.2 |
| N186 | End stage renal disease | 1892 | 278 | 14.7 | 1 | 0.4 |
| E875 | Hyperkalemia | 1845 | 1776 | 96.3 | 500 | 28.2 |
| The 50 most frequently occurring ICD-10-CM codes are presented here; the complete list is available at [https://huggingface.co/datasets/wangyichen25/ICD-10-CM_Validation_Results/viewer]. | | | | | | |
